# The impact of customized head molds on motion and motion-related artifacts from structural and functional MRI scans in children

**DOI:** 10.1101/2021.03.24.21253213

**Authors:** Timothy B. Weng, Ruben D. Vela, Wade Weber, Manwitha Dodla, Anibal S. Heinsfeld, Samuel D. Parker, Blake Simon, Damion V. Demeter, Tehila Nugiel, Lucy Whitmore, Kathryn L. Mills, Jessica A. Church, Michael R. Haberman, R. Cameron Craddock

## Abstract

Although neuroimaging provides powerful tools for assessing brain structure and function, their utility for elucidating mechanisms underlying neuropsychiatric disorders is limited by their sensitivity to head motion. Several publications have shown that standard retrospective motion correction and arduous quality assessment are insufficient to fully remove the deleterious impacts of motion on functional (fMRI) and structural (sMRI) neuroimaging data. These residual errors tend to be correlated with age and clinical diagnosis, resulting in artifactual findings in studies of clinical, developmental, and aging populations. As such there is a continued need to explore and evaluate novel methods for reducing head motion, and their applicability in these populations. Recently, a custom-fitted styrofoam head mold was reported to reduce motion across a range of ages, mostly adolescents, during a resting state fMRI scan.

In the present study, we tested the efficacy of these head molds in a sample exclusively of young children (N = 19; mean age = 7.9 years) including those with ADHD (N = 6). We evaluated the head mold’s impact on head motion, data quality, and analysis results derived from the data. Importantly, we also evaluated whether the head molds were tolerated by our population. We also assessed the extent to which the head mold’s efficacy was related to anxiety levels and ADHD symptoms. In addition to fMRI, we examined the head mold’s impact on sMRI by using a specialized sequence with embedded volumetric navigators (vNAV) to determine head motion during sMRI. We evaluated the head mold’s impact on head motion, data quality, and analysis results derived from the data. Additionally, we conducted acoustic measurements and analyses to determine the extent to which the head mold reduced the noise dosage from the scanner. We found that some individuals benefited while others did not improve significantly. One individual’s sMRI motion was made worse by the head mold. We were unable to identify predictors of the head mold response due to the smaller sample size. The head molds were tolerated well by young children, including those with ADHD, and they provided ample hearing protection. Although the head mold was not a universal solution for reducing head motion and improving data quality, we believe the time and cost required for using the head mold may outweigh the potential loss of data from excessive head motion for developmental studies.

## 1. Introduction

Head motion is perpetually detrimental in neuroimaging research, with even slight movements having the potential to induce artifactual scientific findings (Pardoe et al., 2016; Satterthwaite et al., 2012; Van Dijk et al., 2012). When studying hyperkinetic populations, such as children with ADHD, researchers adhere to stringent criteria for discarding high motion data to reduce the likelihood of reporting false positive findings. These motion criteria can result in wasted data and resources. Unfortunately, even these efforts may not be enough to equalize motion between groups; the mean motion of a hyperkinetic group may still be higher (albeit below an acceptable motion threshold) than the comparison group (Couvy-Duchesne et al., 2016). Applying a threshold for excluding high motion subjects induces a sampling bias when comparing a hyperkinetic group to a comparison group. This sampling bias can result in imaging results being driven by the subsample of individuals that exhibit the least motion who may not be representative of the population (Wylie et al., 2014). Further, since head motion can be highly stereotyped in certain populations and remain stable across multiple scanning sessions (Engelhardt et al., 2017), it can lead to artifactual findings even when the amount of motion does not differ between groups. Consequently, head motion remains an obstacle in neuroimaging studies, particularly in those of developmental disorders.

Although head motion is most commonly discussed in the context of functional MRI (fMRI), it is deleterious to all imaging modalities. In task-based fMRI, head motion can be correlated with the task design, leading to increased false-positive activations (Magland & Childress, 2014). In functional connectivity-based fMRI, head motion inflates correlations between brain areas that are closer together in distance and decreases correlations between areas that are further apart (Power et al., 2012; Satterthwaite et al., 2012; Van Dijk et al., 2012; Yan et al., 2013). Additionally, head motion during diffusion MRI can result in ‘signal dropout’ of entire slices which reduces the amount of data available for analysis and can bias tractography results (Benner et al., 2011; Holdsworth et al., 2012; Kreilkamp et al., 2017; Yendiki et al., 2014). In sMRI, head motion during scan acquisition is negatively correlated with cortical thickness and volume estimates extracted from the scan, even after stringent quality control (Alexander-Bloch et al., 2016; Blumenthal et al., 2002; Pardoe et al., 2016; Reuter et al., 2015). Presumably, head motion blurs the grey matter – white matter interface, making segmentation more difficult, thus influencing volume estimates (Caballero-Gaudes & Reynolds, 2017; Savalia et al., 2017). Furthermore, head motion can lead to artifactual correlations between measures of brain function, brain structure, and phenotype (Ducharme et al., 2016; Siegel et al., 2016).

Due to the prevalence and severity of motion artifacts in MRI studies, many methods have been developed to remove and correct for motion-related artifacts both prospectively and retrospectively. Retrospective motion correction (RMC) entails a series of image processing procedures that are applied to the data after it has been acquired. RMC in multi-slice sMRI data could entail coregistering neighboring slices to correct between-slice in-plane motion, but cannot adequately address through-plane (often head-to-foot) motion, or motion that occurs during the acquisition of a single slice (Godenschweger et al., 2016). For 3D sMRI acquisitions, there is very little opportunity for RMC beyond applying some form of k-space filtering or regularization during image reconstruction aimed at reducing head motion artifacts such as blurring. This form of reconstruction-based RMC can be applied to most imaging modalities, although in most research imaging scenarios k-space data is not easily accessible or saved, making reconstruction-based approaches uncommon. As a result, RMC of sMRI data is limited to discarding motion-contaminated scans identified through careful visual inspection; a motion threshold cannot be determined because motion cannot be objectively measured from 3D sMRI data.

Functional MRI, which is a time series of imaging volumes, is much more amenable to RMC techniques. Motion that occurs between volumes, or between a volume and a reference, can be estimated and nullified using image realignment techniques (Cox, 1996). Additionally, the time series of estimated head motion parameters (x, y, and z translations, pitch, roll, and yaw) can be used to construct a regression model for fitting and subsequently removing motion-induced variations in voxel intensity (Friston et al., 1996). Further RMC can be performed by removing volumes containing motion in excess of a predetermined threshold (Power et al., 2014). Alternatively, other RMC techniques have utilized independent components analysis (ICA) to identify and regress out components that are characteristic of motion (Pruim et al., 2015). This ICA regression-based approach retains the number of volumes that were originally acquired.

Head motion is particularly problematic for diffusion weighted imaging (DWI). Eddy currents generated by quickly switching diffusion gradients on and off will induce geometric distortions in the data that can be exacerbated by head motion. The superposition of these two effects make it difficult to accurately estimate the amount of head motion that occurs during the acquisition, but they can both be addressed using similar realignment methods that are applied to fMRI data. This approach is less effective for sequences that use very high diffusion gradients due to their very low signal-to-noise ratio (SNR) that makes it difficult to calculate an accurate alignment. Head motion estimation can be improved by interleaving volumes collected with no diffusion gradient (b=0) with the higher diffusion gradient images and estimating motion just between these images. This of course will miss out on motion that occurs faster than the time between b=0 volumes. Since DWI is explicitly designed to be sensitive to motion, head motion can be much more insidious for DWI data compared to other imaging modalities. If enough motion occurs at the right time during the acquisition of a DWI slice, the entire brain signal will be dephased by the diffusion gradients, and nothing will be acquired for that slice. This problem can only be addressed during acquisition by preventing head motion or by detecting motion and re-acquiring the missing data.

Although RMC techniques are by far the most commonly performed and most attractive techniques for dealing with head motion, their efficacy is limited by the degree to which the deleterious impacts of motion can be accurately modeled and removed. For example, in fMRI, researchers have demonstrated motion effects that exist after stringent RMC procedures (motion realignment and regression) have been performed (Power et al., 2012; Satterthwaite et al., 2012; Van Dijk et al., 2012; Yan et al., 2013), and similar results have been shown for sMRI <u>(Reuter et al., 2015)</u>. This is also the case for DWI, where head motion can result in missing data for some slices of the brain, or when head motion interacts with the field inhomogeneities to create spatially varying signal dephasing (e.g., around areas of susceptibility) (Le Bihan et al., 2006). Given the limitations of RMC techniques, reducing the impact of head motion on data acquisition or reducing head motion altogether offers an ideal level of protection against head motion artifacts even when RMC methods are available.

Prospective motion correction (PMC) techniques have been developed to measure head motion in real-time and adjust acquisition parameters accordingly to offset the changes in head position (reviewed in Maclaren et al., 2013). For fMRI, this can be done by measuring the displacement between consecutive volumes and repositioning the slice acquisition to nullify any head motion that occured (Thesen et al., 2000). This is attractive since it can be done with only minor modifications to the imaging sequence, but since head motion is estimated from the same data that ultimately becomes the outcome data, it cannot completely remove all of the motion that occurs. For example, in this scenario motion between the first and second TR is used to estimate motion that is used to correct the acquisition of the third TR. This correction does not remove the motion between the first and second TR, and assumes that the motion between the second and third TR is consistent with the motion between the first and second, which may not be the case (Thesen et al., 2000; White et al., 2010). This problem can be fixed by using a separate source of information for estimating head motion, such as another imaging sequence interleaved with the first that measures the head (PROMO; White et al., 2010) or a MRI fiducial placed on the head (Muraskin et al., 2013), or an external optical based head tracking system (Maclaren et al., 2012; M. Zaitsev et al., 2006). This approach has been used effectively with a variety of imaging sequences and is popular for structural MRI (e.g., PROMO; White et al., 2010). In one approach, volumetric navigators (quickly acquired low resolution volumes of the brain) are embedded into the dead spaces that occur in commonly used sMRI sequences. Head motion can be measured between these navigators, in the same way it is measured between consecutive fMRI volumes, and then used to estimate motion that is nullified by changing the proscription of the next acquired segment of k-space (Tisdall et al., 2012). While these PMC approaches show promise, they have a few drawbacks of their own. MRI navigators require sequence customizations that may not be possible for all imaging modalities. Optical systems require additional equipment and line of site to the participant’s head, which may not be feasible. Also optical systems measure skull motion, which may be very different from brain motion (Badachhape et al., 2018). Finally, since head motion estimates are used to change the image acquisition proscription, any error in this estimate can result in over-corrections that introduce motion into otherwise low-motion data and the uncorrected data cannot be recovered (Godenschweger et al., 2016).

There are MRI sequence read-out schemes with reduced sensitivity to motion that have been proposed for fMRI that can also be classified as PMC. These include 3D stack of spirals (Hu & Glover, 2007), PRESTO (Liu et al., 1993), echo volumar imaging (van der Zwaag et al., 2006). Similarly, there are established sequences with reduced motion sensitivity that have been used extensively in abdominal and cardiac imaging (e.g., radial MRI). However, these methods have not taken hold in neuroimaging either due to poor spatial or temporal resolution, increased SNR, or perhaps other reasons. Therefore, these methods are not considered further here.

Given the limitations of RMC and PMC techniques, the best method to limit the deleterious impacts of motion on neuroimaging data is to prevent motion from happening in the first place. Behavioral methods and physical restraints have been developed to do just that. Real-time systems have been developed that estimate head motion while the data are being acquired. This information can be displayed to the subject during scanning to have them self-regulate their motion (Greene et al., 2018; Yang et al., 2005), or it can be displayed to the operator who can counsel the subject or restart or extend high motion scans (Cox & Jesmanowicz, 1999; Dosenbach et al., 2017; Esper et al., 2019). Real-time head motion feedback can require modifications to the MR system hardware and may need to use navigated sequences (e.g., vNAV) or external tracking devices to track motion during non-fMRI scans. These real-time approaches have shown promise at reducing head motion, although providing motion feedback directly to the subject during fMRI may interfere with the primary task being performed (Greene et al., 2018; Yang et al., 2005).

One source of head motion during scanning could be the subject’s anxiety or discomfort during the scanning procedure. This can be ameliorated using one or more mock scanning sessions, during which the subject is led through a simulated MRI scan that uses a reasonable facsimile of a MRI system and includes listening to pre-recorded sounds of various scanning sequences. A variety of different interventions can be included in this process to acclimate the subject to scanning and train them to keep their head still during scanning (Simhal et al., 2021). Mock scanning in children can improve scan quality and reduce head motion, including those with ADHD (de Bie et al., 2010; Epstein et al., 2007; Pua et al., 2020). Strategies for improving comfort and reducing anxiety during actual scanning include covering the subject in a weighted blanket (Horien et al., 2020), allowing them to watch an engaging video (Alexander et al., 2017; Greene et al., 2018; Vanderwal et al., 2015), and aromatherapy (Davis, 2016; Schellhammer et al., 2013). For example, in a sample of children, including those with autism spectrum disorder, (mean age = 11.2 years), Horien et al. (2020) achieved high-quality, low-motion fMRI data by using a combination of mock scanning with formal instruction, weighted blankets, and an incentive system.

A few different methods have been proposed to physically restrain head movement. Bite bars fitted with custom dental impressions have been reported to perform well at reducing head motion (Menon et al., 1997), but their use is not common, perhaps due to concerns over subject discomfort (Zaitsev et al., 2015). A less aggressive and more commonly used method for limiting head motion is to use foam padding that expands and fills gaps between the head and MRI coil, but this has been reported to be insufficient in reducing motion (e.g., Reuter et al., 2015).

Recently, precision-fitted head molds were reported to reduce head motion in resting state fMRI data from a sample ranging from 7 to 28 years old (Power et al., 2019). Using optical imaging, participants’ head shapes were scanned and then milled into Styrofoam head molds by a CNC machine (https://caseforge.co/). The insides of the molds were milled to fit each participant’s head, and the outsides of the mold were shaped to fit the MRI coil. Thus, the head molds more precisely restricted head motion from occurring compared to conventional methods. Compared with most PMC techniques, head molds can be used with any imaging sequence without impacting image quality, imaging parameters, or scan length. The customized styrofoam head molds were reported to provide comfort while restraining head motion (Power et al., 2019). Therefore, head molds are an attractive solution compared to the many varieties of methods for preventing head motion or correcting for its impact on imaging data.

In nearly all of the participants, Power et al. (2019) found the head mold reduced head motion of varying magnitudes. Indeed, the reductions in head motion translated to diminished motion-related artifacts on functional connectivity calculations. In another study, Jolly et al. (2020) compared motion parameters between a group of young adults wearing the head mold to another group without the head mold. Functional scans were acquired from both groups while they watched a movie and performed a spoken recall task following the movie. However, Jolly et al. (2020) found no significant differences in head motion between the groups.

Power et al. (2019) and Jolly et al. (2020) investigated head mold effects on functional MRI; however, it remains unknown whether these molds are effective for reducing head motion during sMRI and improving sMRI data quality and subsequent estimates of cortical thickness and volume. Additionally, the majority of the participants studied by Power et al. (2019) were over the age of 10 years (mean age = 15 years), and Jolly et al. (2020)’s study was conducted on undergraduate and graduate students (mean age = 20.8 years). Therefore, as discussed by Horien et al. (2020), it is not known how the head molds perform in an exclusively younger population, especially one including participants with developmental disorders.

In the current study, we build upon the works of Power et al. (2019) and Jolly et al. (2020) by assessing the efficacy of head molds for ameliorating head motion and its effects on image artifacts in an exclusively young sample (ages 5 - 10 years), including a subsample of children with ADHD. We extend the work of Horien et al. (2020) by testing whether head molds and movie watching reduce head motion beyond what can be accomplished using mock scanning and a weighted blanket.

Each participant completed a dedicated session for mock scanning before we acquired two sets of sMRI and fMRI scans. One scan set was collected using the head mold and the other without, and the scans were counterbalanced in order across participants. Using a specialized sMRI sequence embedded with vNAVs, we directly measured head motion during sMRI in order to more accurately evaluate the head mold’s impact on structural metrics. To evaluate the feasibility and tolerability of the head mold, we concluded the scanning sessions with a brief interview that gauged participants’ scanning experience and tolerance of the head mold. We investigated whether head molds would reduce head motion compared to standard scanning procedures and whether those reductions would result in improved data quality. We also tested the extent to which reductions in head motion impacted estimations of cortical volume and functional connectivity. Prior to data collection, we evaluated the noise dosage levels of the head mold condition, given the limited space available for additional hearing protection devices in the head coil while the head mold is on. We also assessed whether the head molds could balance head motion between typical and ADHD populations, and whether clinical variables such as ADHD diagnosis and trait anxiety, as determined by parent-reported dimensional measures, can predict head mold efficacy.

## 2. Materials and Methods

### 2.1 Participants

We enrolled seven children with an ADHD diagnosis (ADHD), and 14 typically-developing controls (TDC), as determined by parent self-report. Recruitment was limited to children aged 5-10.99 years old. The parents of all participants reported no MRI contraindications and no history of head injuries or seizures. There were six sibling pairs enrolled into the study; in five pairs both siblings were TDC and in one pair both siblings had ADHD. Two children (one TDC male and one ADHD male) were excluded from the analyses because they did not successfully complete MRI scanning. One of the excluded participant’s head mold did not fit on the day of the scan, possibly due to the 11-week delay between ordering the mold and the scanning visit (the scanning visit was delayed due to technical issues with the scanner). The other excluded participant experienced anxiety related to the movie (chosen by the child with their parent’s permission) shown during the scan and they asked to terminate the study. The final analyses included data from 13 TDC (3 males, 10 females, age = 7.5 ± 1.5 years) and six ADHD (3 males, 3 females, age = 8.7 ± 1.4 years) participants. The study protocol was approved by The University of Texas at Austin Institutional Review Board (IRB).

### 2.2 Noise Safety

One disadvantage of customized head molds is that they limit the types of audio delivery systems and hearing protection that can be used. Our imaging center provides three systems for audio delivery: Optoacoustics OptoActive active noise canceling headphones (Optoacoustics Ltd., Mazor, Israel), Sensimetrics Model S14 insert earphones (Sensimetrics Corporation, Gloucester, MA), and the Siemens pneumatic headphones that come standard with the Skyra system (Siemens Healthcare, Malvern, PA). To ensure adequate hearing protection, our lab procedures are to use earplugs under the Optoacoustic or pneumatic headphones with over-the-ear earmuffs for additional hearing protection when using the Sensimetrics earphones. This double-protection mitigates the variability in hearing protection afforded by earplugs due to incorrect insertion (Murphy et al., 2011). However, neither of these configurations are possible when using head molds. Instead, we were limited to using the Sensimetrics earphones with limited acoustic foam padding placed over the ear. Previous literature has shown that a similar configuration of in-ear headphones, foam padding and a loosely-fitted foam helmet can substantially reduce MRI noise experienced by participants (Ravicz & Melcher, 2001).

Following Ravicz & Melcher (2001), we performed a series of experiments prior to data collection in order to verify the quality of hearing protection achieved by our specific configuration (Caseforge head mold in combination with the S14 in-ear monitors and foam padding). In the first experiment we recorded the sound produced by the Siemens Skyra 3T MRI system located at the Biomedical Imaging Center at The University of Texas at Austin while running standard functional and structural MRI sequences (same sequences and parameters as used for data collection below). A standard cylindrical water phantom was placed in the head coil and was moved to the center of the bore to provide signal for pre-scanning frequency and shim adjustments (without which these adjustments would not complete and scanning would not occur). Sound was recorded using an MRI-compatible Optimic 1155 fiber optic microphone placed on the phantom at the center of the bore (Optoacoustics Ltd., Mazor, Israel). The voltage output from the microphone was calibrated to output pressure values using a Larson Davis CAL2000 precision acoustic calibrator (Larson Davis, Depew, NY). This calibration was validated using reference measurements collected in an acoustic anechoic chamber at The University of Texas at Austin’s main campus.

The second experiment involved a series of measurements collected in the anechoic chamber aimed at estimating hearing protection properties for the various combinations of the head mold, ear plugs, and foam padding (Ravicz and Melcher, 2001). Each trial of the experiment involved a 15-second duration exponential chirp ranging from 20 - 20948 Hz being played from a self-powered loudspeaker (ADAM Audio AX7 Active Nearfield Monitor, Berlin, Germany) and recorded using two GRAS 40AC ½” externally-polarized free field capsule microphones (GRAS Sound and Vibration, Holte, Denmark). One microphone was mounted in the left ear of a Knowles Electronic Manikin for Acoustic Research (KEMAR; GRAS Sound and Vibration, Holte, Denmark) (Figure S1) located approximately 1.5 meters from the loudspeaker, and the other was suspended above the KEMAR to provide a free-field reference (Figure S2). The experiment was repeated with several different configurations of hearing protection applied to the KEMAR: (1) no hearing protection, (2) earplugs only, (3) foam inserts only, (4) head mold only, (5) earplugs and foam, (6) earplugs and head mold, (7) foam and head mold, and (8) earplugs, foam, and head mold. A custom head mold was manufactured for the KEMAR using the procedures described in Section 2.3.

The measurements collected from the two microphones were compared within and between each of these conditions to estimate insertion loss (IL) and noise reduction (NR). IL is the ratio of the sound level measured by the microphone mounted in KEMAR for one configuration of hearing protection to the sound level measured in the reference configuration of no hearing protection. NR is the ratio of the sound pressure measured by the free field microphone to the sound pressure measured by the microphone in KEMAR in each configuration. Using NR, the head-related transfer function (HRTF) is determined, which is a model of how a sound transmitted from a sound source is transformed by space and any other intervening materials (i.e., hearing protection) before being received by the ear. Mathematically, the sound received at the KEMAR ear can be calculated by convolving the HTRF with the input time signal in the time domain or by multiplying the frequency specific coefficients of the HRTF by the corresponding amplitudes of the input signal’s frequency components in the frequency domain. Using this strategy, the HRTF for each configuration was applied to the sounds recorded during MRI scanning in the first experiment to estimate the extent of hearing protection that they provide.

### 2.3 Mock Scan and Head Mold Production

In the first of two visits, parents and children met with research staff to be informed about study procedures, complete a mock scan, provide written consent, and to collect a 3D scan of their head and face for head mold construction. The mock MRI was performed prior to obtaining consent to acclimate the children to the MRI scanning equipment and procedures. The mock system is built from the external enclosure from a decommissioned system. The various components of the MRI system (e.g., bore, head coil, hearing protection) were explained to the children and they were given the opportunity to explore each item. Once they were comfortable with the system they were asked to lay on the scanner bed, hearing protection ear muffs were placed over their ears, a mock head coil was placed over their head, and they were moved into the scanner bore. While in the bore they were asked to practice laying still for one minute while listening to scanner noise played through speakers placed next to the scanner bore. All 21 enrolled participants were comfortable with this procedure and willingly provided informed written consent (consent from the parent, assent from the participant).

After providing consent, a 3D optical scan was collected of each participant’s head and face using a hand-held camera provided by the Caseforge company (https://caseforge.co). A swim cap and diving hood was placed over the participant’s head to flatten and smooth the shape of the hair and to keep it out of their face. Collected 3D scans were transmitted to the Caseforge company where they were used to manufacture a customized Styrofoam head mold and shipped a few days later. The insides of the molds were milled to closely fit each participant’s head, and the outside of the molds were shaped to fit the MRI coil, thus providing head restraint.

### 2.4 MRI Scanning

During the second visit, each participant completed MRI scanning with (MOLD+) and without (MOLD-) the head mold, counterbalanced in order across participants. In both conditions we used MR-compatible insert earphones fitted with foam canal tips that expand inside participants’ ear canals (Model S14, Sensimetrics, Woburn, MA) for communicating to participants, delivering auditory stimuli, and hearing protection. During MOLD+ conditions, small pads of sound dampening foam were placed over the participants ears, underneath the head mold. In MOLD-conditions, over-the-ear headphones were used, and foam pads were placed between the headphones and the headcoil and between the forehead and the headcoil. This is our standard practice for providing hearing protection and limiting head movement to the best of our ability. Participants were also covered with a child-size weighted blanket (6 lbs) in order to help reduce anxiety and further assist with motion reduction. This is especially beneficial for those with neurodevelopmental disorders such as ADHD and autism spectrum disorder (Horien et al., 2020; Nordahl et al., 2016).

Scanning was performed on a Siemens MAGNETOM Skyra 3T MRI scanner at The University of Texas at Austin Biomedical Imaging Center (BIC). Each participant completed two runs of 13.5 minute scan protocol for a total scan time of 27 minutes. Each run consisted of a high-resolution structural image and a functional image. Both runs began with localizer scans and a short vNAV initialization scan. During all scans, children were shown a movie that they and their parents selected from Netflix Kids. Film watching reduces head motion during MRI scanning in young children (Alexander et al., 2017; Greene et al., 2018; Vanderwal et al., 2015). After the first run of scans, participants were given a break and allowed to move or stretch for a few minutes. Then, the scanning configuration was changed depending on the counterbalance assignment (i.e., head mold or earphones added or removed), and the child was returned into the scanner bore to complete the second run. Participants were reminded to lay still and relaxed between each scan and were given ample time to adjust their position in order to lay comfortably during scanning.

The structural images were acquired using a volume-navigated (vNAV) T1-weighted MPRAGE sequence (TR = 2.5 s, TE = 2.9 ms, inversion time = 1.09 s, flip angle = 8°, 256 × 256 matrix, 1 mm isotropic resolution) (Tisdall et al., 2012). This sequence incorporates low-resolution 3D navigators into the MPRAGE acquisition to estimate head motion as the root mean square deviation of head displacement per minute (RMSpm). Although this sequence has the capability to modify its acquisition in real-time to correct for motion, this functionality was not used during our acquisitions. The functional images were obtained using a T2*-weighted echo-planar imaging (EPI) sequence (TR = 2 s, TE = 32.2 ms, 256 × 256 matrix, flip angle = 71°, 2.5 mm isotropic resolution, scan duration = 6 min).

### 2.5 Correcting Head Mold Fit

For some participants the head mold was too tight or uncomfortable when attaching the MR head coil. In these cases (N = 9), we modified the outside of the head mold using a utility knife to improve its fit. The most common modifications were removing material from next to the nose and shaving down the material over the forehead.

### 2.6 Subjective Measures

Parents of participants completed two computerized questionnaires, the Screen for Child Anxiety Related Emotional Disorders (SCARED) and Strength and Weaknesses of Attention-Deficit/Hyperactivity-symptoms and Normal-behaviors (SWAN) using REDCap (Research Electronic Data Capture; Harris et al., 2009, 2019). The SCARED is a 41-item self-report or parent-report measure of anxiety symptoms in children and is useful for identifying markers of generalized anxiety, social anxiety, separation anxiety, and school avoidance. We chose to administer the parent-report version due to the young age of this participant sample. The SWAN uses observer ratings to help assess symptoms of ADHD in children, providing separate dimensional measures of inattention and hyperactivity/impulsivity. The parents of sixteen (14 TDC, 2 ADHD) participants completed the assessment remotely after their second visit, the remaining five participants (all ADHD) completed the assessment during their enrollment visit.

After the MRI scans, participants were asked to provide feedback on their subjective experience during each scanning run. This included responding to a 5-point version of the Wong-Baker faces scale (Wong-Baker FACES Foundation, 2020) to report how they felt during the scan overall and during the scan while wearing the head mold. In addition, they were given 4 free-form response questions: ‘*Which did you like better, the scans with the helmet or without?’, ‘What was the best part of the scan?’, ‘What was the worst part of the scan?’, ‘Would you do another scan with the helmet?’*.

### 2.7 Image Preprocessing and Analysis

Following Reuter et al. (2015), we tested whether the head mold-related changes in motion would influence brain volume estimates from a variety of tools. FSL Siena (Smith et al., 2002) estimated the percent brain volume change between MOLD- and MOLD+ conditions using the two-time-point estimation function. The Computational Anatomy Toolbox 12 (CAT12) longitudinal processing running in SPM12 on Matlab R2019a was used to calculate volume based morphometry (VBM) estimates of total brain, gray matter, and white matter (Gaser & Dahnke, 2016). CAT12 also provided an index of image quality. The FreeSurfer 6 Longitudinal Pipeline was used to estimate cortical thickness and gray matter volume (Fischl et al., 1999a, Fischl et al., 1999b, Reuter et al., 2012).

Imaging data were minimally preprocessed using Configurable Pipeline for the Analysis of Connectomes (C-PAC). The structural data were skull-stripped using HD-BET (Isensee et al., 2019). Skull-stripped images were then used as inputs to CAT12, FreeSurfer, and FSL Siena for brain volume estimation. Functional preprocessing in C-PAC involved motion correction, transformation to MNI template, and resampling to 3 mm isotropic resolution.

For the functional data, the primary summary metric of participant motion used was mean framewise displacement (FD_Power_) using head position estimates from *3dvolreg*. For the structural data, real-time estimates of head motion were extracted from the vNav images in the form of average displacement per minute (Motion_RMS_ mm/min).

Functional connectivity was measured as Pearson’s correlation between pairs of regions from the 264 regions of interest (ROIs) atlas (Power et al., 2011). First, the timeseries for each ROI was extracted from the preprocessed data and detrended. Then, for all possible ROI pairs, the time series were correlated, yielding a 264 x 264 correlation matrix for each scan. Another 264 x 264 matrix was created containing the Euclidean distance between every ROI pair. These matrices served as the basis for assessing whether head molds impacted distance-dependent motion artifacts in functional connectivity analyses.

## 3. Results

### 3.1 Hearing Protection Assessment

Figures S3 and S4 illustrate the impact of the different combinations of hearing protection tested on the frequency content of the sound generated by the structural and functional MRI scanning sequences. All combinations of hearing protection involving the ear plugs reduced the noise level across the spectrum, with the addition of the head mold and foam padding providing greater noise reduction at higher frequencies than what is afforded by ear plugs alone. Table S1 contains dosage levels for different hearing protection combinations at different scan lengths of structural and functional MRIs. Given that noise dose is additive, the sound exposure for a scanning session consisting of a 7.5-minute sMRI and 45 minutes of fMRI without any hearing protection is 356% of the limits specified by OSHA regulations for an 8-hour day (OSHA, 2008). The head mold alone reduced the exposure to 124% of the limit, which exceeds OSHA regulations. Using the head mold in combination with earplugs or with earplugs and additional foam padding reduced the noise exposure to 2.5% and 1% of the 8-hour exposure limit, respectively. Both of these are more than adequate, but we chose to use the foam padding in all of our scanning as it helped to secure the earplugs in place.

**Table 1.** Descriptive statistics for participants who completed scanning. Values are reported as mean (standard deviation).

|  | TDC | ADHD |
| --- | --- | --- |
| N | 13 | 6 |
| Age | 7.54 (1.51) | 8.67 (1.51) |
| ADHD Hyperactive/Impulsive Score | 0.75 (1.29) | 4.50 (3.56) |
| ADHD Inattentive score | 1.00 (1.41) | 6.67 (1.21) |
| Generalized Anxiety | 2.75 (2.63) | 4.83 (5.42) |
| Panic | 1.25 (0.87) | 2.83 (4.67) |
| School Avoidance | 0.08 (0.29) | 0.00 (0.00) |
| Separation Anxiety | 1.75 (2.01) | 3.00 (3.16) |
| Social Anxiety | 3.42 (3.15) | 6.17 (2.93) |
| Total Anxiety | 8.54 (7.05) | 16.83 (14.97) |

### 3.2 SWAN/SCARED Assessment

Parent-reported SWAN and SCARED questionnaires were completed for 12 of the 13 TDC children who completed MRI scanning. As expected, none of these 12 children met criteria for ADHD as measured by the SWAN rating scale. Results from the SCARED revealed that of this group, one child met criteria for Separation Anxiety Disorder, another met criteria for Social Anxiety Disorder, and another for both Separation and Social Anxiety Disorders. SCARED and SWAN assessments were not completed by the parents of the TDC child who did not complete the MRI scan.

Of the six children with ADHD who completed scanning, results from the SWAN revealed that two met criteria for inattentive type (ADHD-PI), and three met criteria for combined type ADHD (inattentive and hyperactive/impulsive; ADHD-C). One of the children with ADHD was one point subthreshold for ADHI-PI. Results from the SCARED revealed that two met criteria for potential Social Anxiety Disorder and one met criteria for Anxiety Disorder, Panic Disorder, Generalized Anxiety, Separation Anxiety and Social Anxiety Disorders. The child with ADHD who did not complete MRI scanning met criteria for combined type ADHD and School Avoidance.

### 3.3 Tolerance

A Wilcoxon Rank Sum Test indicated that there were no significant differences in the distribution of tolerability ratings between the two scanning conditions (W = 18, *p* > 0.05) (Fig. 1A). However, children were less positive about the MOLD+ condition (74% of responses for the MOLD-condition were positive, compared to 42% of responses for the MOLD+ condition). This reduced enthusiasm was reflected in more neutral ratings for MOLD+ (26% of responses for the MOLD-condition were neutral, compared to 57% in the MOLD+ condition). One child indicated a negative response for the MOLD+ condition (Sad/Neutral in Fig. 1A). Despite this reduced enthusiasm for the head mold, only 52.6% of the participants preferred the MOLD-scans (Fig. 1B) and 5.3% of the participants indicated they would not return for another scan using the head mold (Fig. 1C).

**Fig. 1.**
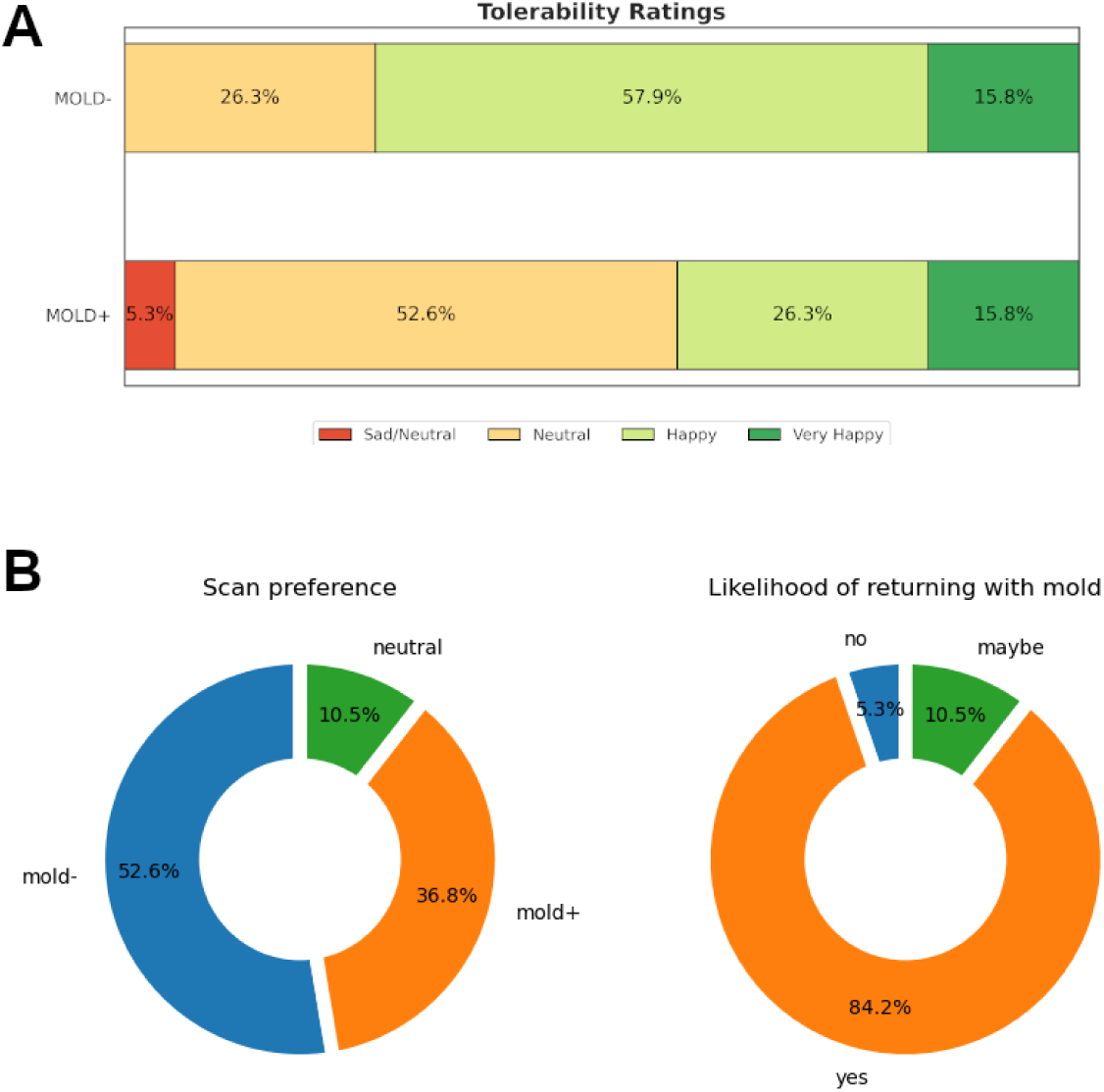
Tolerability of head molds. **A.** Diverging stacked bar charts showing the distribution of responses from a 5-point graphical Likert scale (1 = Very sad face; 2 = Sad face; 3 = Neutral face; 4 = Happy face; 5 = Very happy face). The charts illustrate that 74% of responses for the MOLD-condition were positive, whereas 42% of responses for the MOLD+ condition were positive. 26% of responses for the MOLD-condition were neutral, compared to 57% in the MOLD+ condition. *Note*: One child responded they were between a sad and neutral face (sad/neutral). **B.** Pie chart comparing the proportions of participants who preferred the one scan condition over the other. **C.** Proportion of participants who indicated their willingness to return for another scan with the head mold on.

### 3.4 Head Motion and Image Quality Assessment

Head motion during the functional scans has been used as a proxy for motion during structural scans (Pardoe et al., 2016; Savalia et al., 2017) but has not been directly validated given the lack of estimates of head motion from structural MRI data. We tested whether fMRI head motion is suitable as a surrogate for sMRI head motion using Pearson’s correlation to measure their linear relationship and Lin’s concordance correlation coefficient (ρ_c_) to measure their equivalence (Lin, 1989). Root means square deviation estimates of motion (Jenkinson et al., 2002) were first normalized by repetition time (TR) to correct for differences in timing between sMRI and fMRI sequences. Correlation values were calculated after excluding the outlier denoted by the x symbol in Figure 2. With the head mold off (MOLD-), the Lin’s concordance correlation coefficient (ρ_c_) between the functional and structural scans was 0.14 (95% CI: −0.18 - 0.43), and the Pearson’s correlation coefficient (r) was 0.82. With the head mold on (MOLD+), there was relatively higher concordance (ρ_c_ = 0.42; 95% CI: 0.12 - 0.65) compared to MOLD-, whereas the Peason’s correlation coefficient was lower (r = 0.57).

**Fig. 2.**
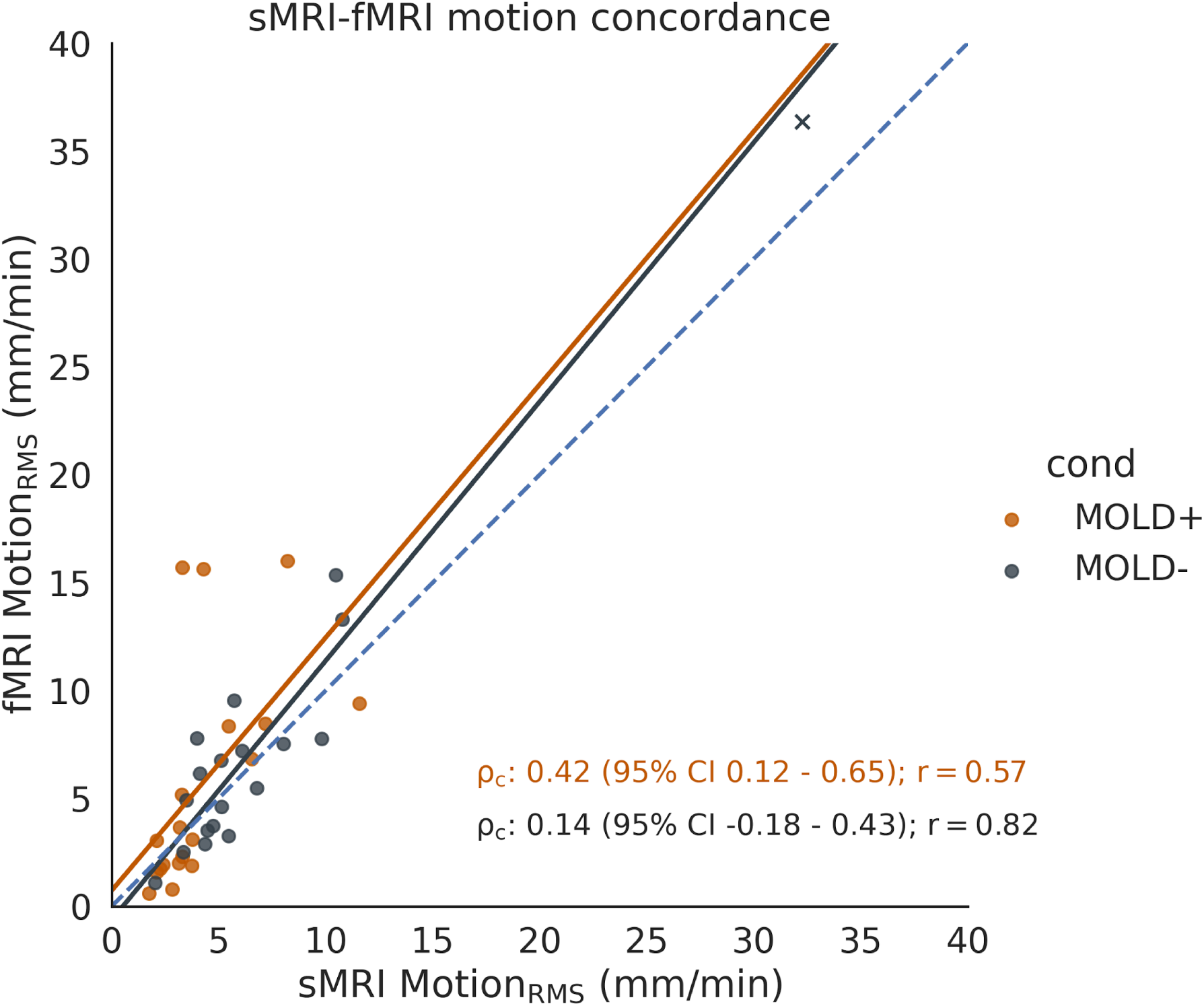
Correspondence of head motion between the functional and structural scans. Solid lines represent a linear regression. Note that the regression lines do not correspond to Lin’s concordance correlation coefficient (⍴_c_). Dashed line represents the line of perfect concordance (⍴_c_ = 1).

#### 3.4.1 Functional

We assessed the impact of the head mold on head motion during the functional scan using mean framewise displacement (mean FD_Power_) as defined by Power et al. (2012) to be consistent with other evaluations of the head mold (Jolly et al., 2020, Power et al., 2019). Additionally, we tested whether the head mold reduced the number of large spikes in head motion, as they are known to be particularly problematic for fMRI analyses. We did this by testing whether the head mold reduced the proportion of scans with FD_Power_ greater than 0.3 mm, which was the threshold for motion scrubbing used in Power et al., (2019). Across the sample, the head mold did not significantly reduce mean FD_Power_ (*t*(18) = −1.83, *p* = 0.084). However, the proportion of scans with FD_Power_ above 0.3 mm was significantly reduced by the head mold (*t*(18) = −2.64, *p* < 0.05).

The lines depicting participant-level changes in Figure 3 reveal a variability of responses. We sought to characterize the variability in motion reductions by examining how head mold-related changes in motion translated to changes in data quality. Using a QC rating threshold of mean FD_Power_ < 0.2 mm (Ciric et al., 2017), we identified changes in QC ratings between MOLD- and MOLD+ (Figure 3)^1^. Note that for our scan parameters, this FD threshold is equivalent to 0.1 mm displacement per second, compared to the 0.11 mm/sec threshold used by Ciric et al. (2017). We found seven participants (5 TDC, 2 with ADHD) whose scans failed QC without the head mold but passed QC with the head mold (fail - pass). Additionally, eight individuals (4 TDC, 4 with ADHD) had unusable data for both conditions (fail - fail). There were four TDCs whose data quality passed both with and without the head mold (pass - pass). Finally, none of the MOLD-scans that passed QC failed QC during the MOLD+ condition (pass - fail). Figure 4 shows example motion traces for each of the three cases of QC rating changes. Across all individuals, the head mold increased the percentage of total usable scans from 21% to 58% adhering to a QC threshold of mean FD_Power_ < 0.2 mm (Figure 5). The same results were present when examining the percentage of scans with FD greater than 0.3 mm. For this QC metric, we used a threshold of 20% (30 volumes, or ∼5 minutes of remaining sub-threshold data), given a desire to retain at least 5 minutes of data (Van Dijk et al., 2010).

**Fig. 3.**
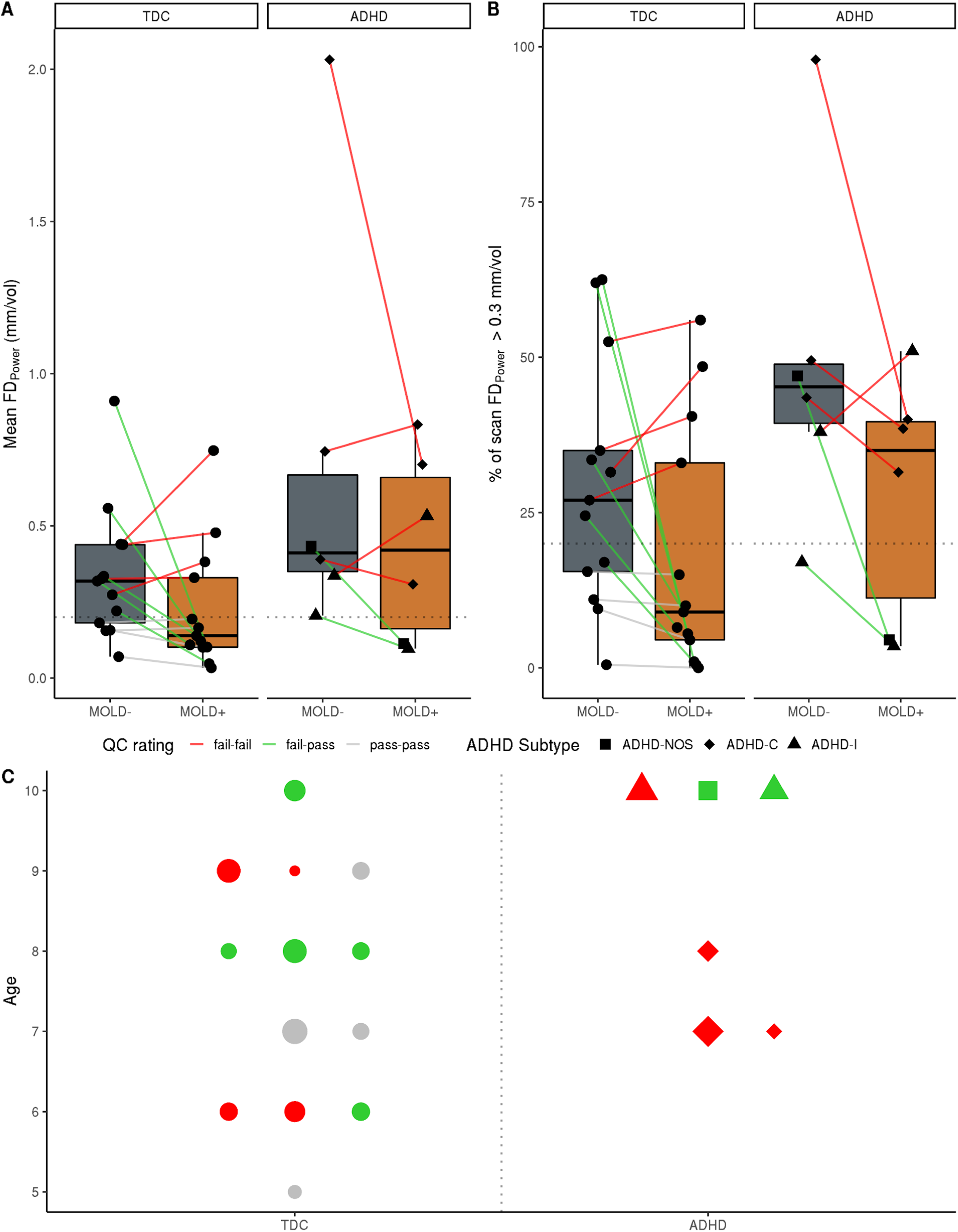
Characterizing head motion during fMRI scan and its impact on data quality. **A.** Mean framewise displacement (FD_Power_) with and without the head mold. **B.** Percentage of the scan with FD > 0.3 mm. For both **A** and **B**, each colored line resembles within-participant change between MOLD- and MOLD+. The colors of the lines indicate how the data quality was affected by the head mold at a given QC rating threshold. The horizontal dotted line denotes the QC threshold used for defining data quality (Mean FD_Power_ < 0.2 mm; 20% of scan with FD greater than 0.3 mm). **C.** Head mold-related impacts on data quality as a function of age, ADHD diagnosis, and anxiety. As in the plots in **A** and **B**, each marker resembles individual participants, and the colors denote the changes in QC ratings between MOLD- and MOLD+. The size of the symbols denotes the total anxiety score from SCARED.

**Fig. 4.**
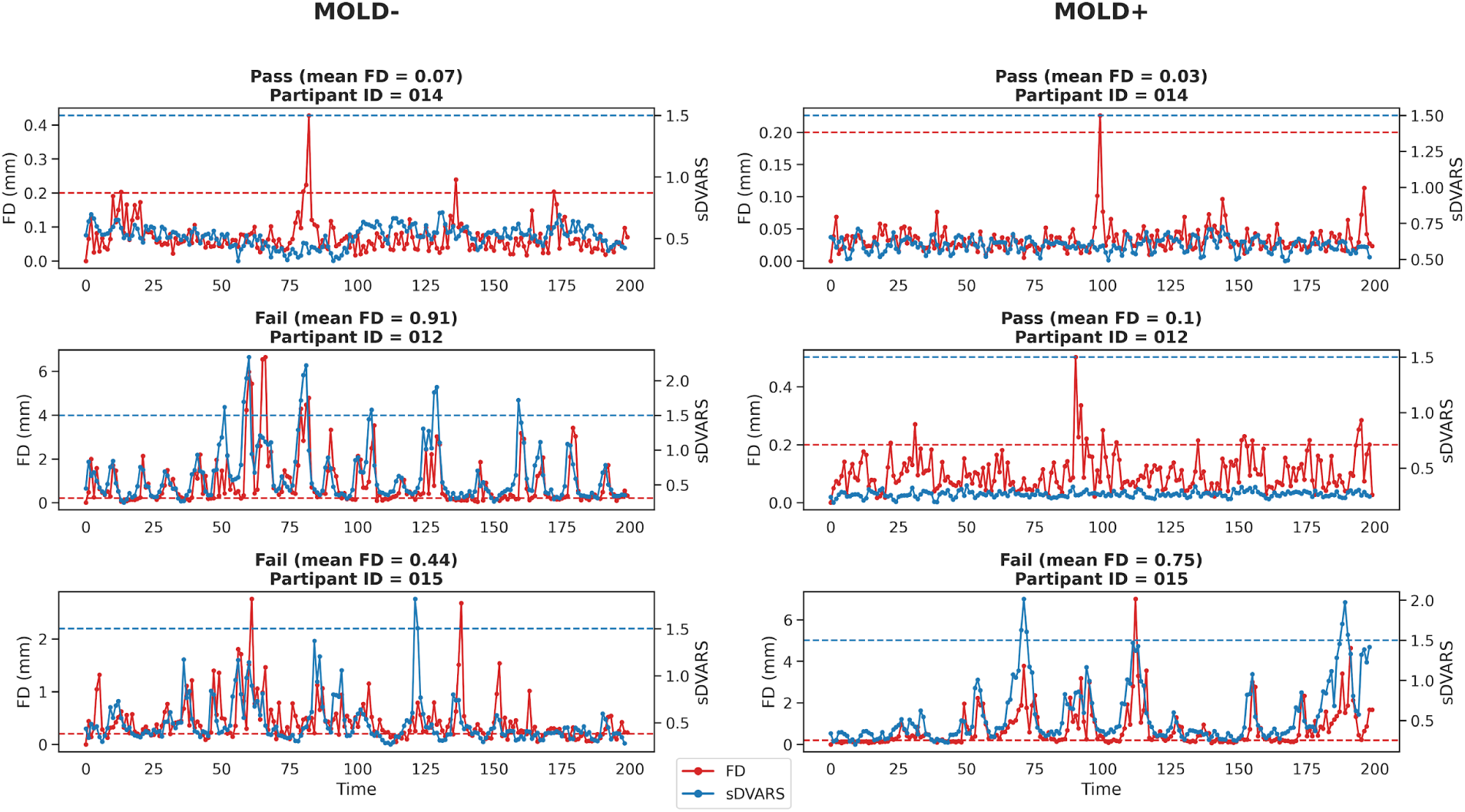
Example motion traces for scans and their QC ratings. Each row represents a single participant. In all plots, the red trace follows framewise displacement (FD) and the blue trace follows DVARS. Dotted horizontal lines represent QC thresholds (FD < 0.2 mm, sDVARS < 1.5). *Top:* This is an example where mean FD was below the QC threshold (0.2 mm) for both scan conditions. *Middle:* This trace shows how the head mold reduced FD. *Bottom*: This is an example where the head mold did not make a difference in head motion. All 3 example participants were TDC with low anxiety.

**Fig. 5.**
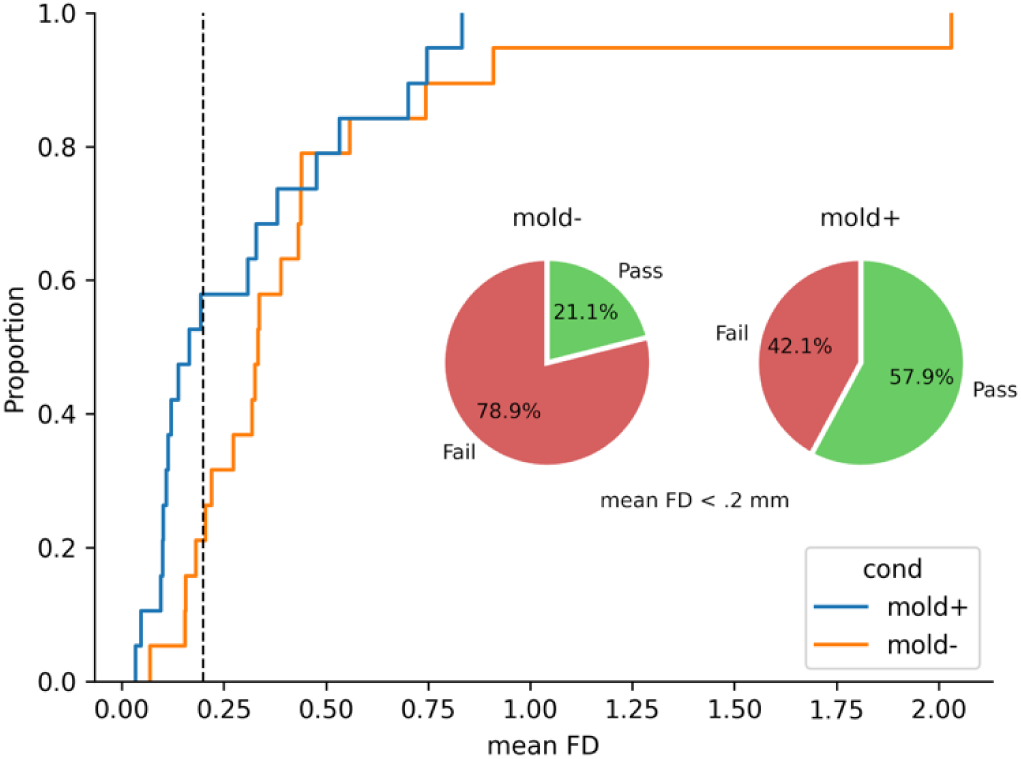
A cumulative distribution function showing that MOLD+ scans have a higher probability of having a lower mean FD compared to MOLD-scans. At mean FD_Power_ = 0.2 mm (vertical dashed line in black), a commonly used threshold for QC ratings, the head mold increased the percentage of usable scans from 21% to 58%.

Given the variability of head mold responses, we sought to determine whether age, anxiety, and ADHD diagnosis had any predictive value on the head mold-related changes in motion. Without the head mold (MOLD-), mean FD_Power_ was greater in ADHD than TDC group, although the results did not reach statistical significance (*t*(18) = 1.73, *p* = 0.10). Additionally, mean FD_Power_ was not correlated with age (r = −0.06) or total anxiety score (r = −0.17). Although there were too few observations to test the statistical significance of subtype differences, head motion appears worse for ADHD-C subtype (N = 3) than ADHD-I subtype (N = 2). None of the data from children with ADHD passed the mean FD quality threshold in the MOLD-condition. Using the head mold (MOLD+) improved head motion in four out of six of the children with ADHD, but data from only two of the six passed the mean FD quality threshold in the MOLD+ condition. The head mold was not able to sufficiently improve functional data in any of the children with combined subtype and one of the two children with inattentive subtype. Similar to MOLD-, mean FD_Power_ during MOLD+ trended greater in ADHD compared to the TDC group (*t*(18) = 1.72, *p* = 0.10). There was no significant relationship between total anxiety score and the success of the head mold for rescuing data, although the head mold did not reduce motion in any of the younger ADHD participants.

#### 3.4.2 Structural

The head mold tended to decrease motion during the structural scan, as indexed by Motion_RMS_, although the results were not statistically significant across the sample (*t*(18) = 1.92, *p* = 0.07). The head mold decreased head movement in 12 participants, increased movement in two participants, and had little to no impact on movement for five participants.

We then examined whether the changes in head motion impacted the quality of the sMRI scans. Currently, qualitative assessments are the primary method used for assessing sMRI quality and identifying scans to exclude. Accordingly, trained raters (co-authors DD, TN, LW, KM, and JC) rated defaced images on a 5-point rating system that was based on the pervasiveness of hallmark motion artifacts such as blurring and ringing (Pardoe et al., 2016). The raters were blinded to the scan condition and the order of scans were randomly shuffled among the raters. Inter-rater reliability was excellent (ICC = 0.94). Given the high inter-rater reliability, the mode score was chosen to represent the quality of each scan.

Indeed, scans that were rated poorly contained higher head motion compared to scans that were rated as higher quality (Spearman’s ρ = −0.81, p < 0.001; Figure 6). This finding verified that head motion influences data quality based on visual inspection. Ratings were then grouped into the following categories: 1 and 2 = Fail; 3 = Warn, 4 and 5 = Pass. Then, a boundary was established between the upper end of pass ratings and the lower end of fail ratings (Motion_RMS_ = 5.29 mm/min). Based on this threshold, images labeled as Warn were then allocated to Pass and Fail labels.

**Fig. 6.**
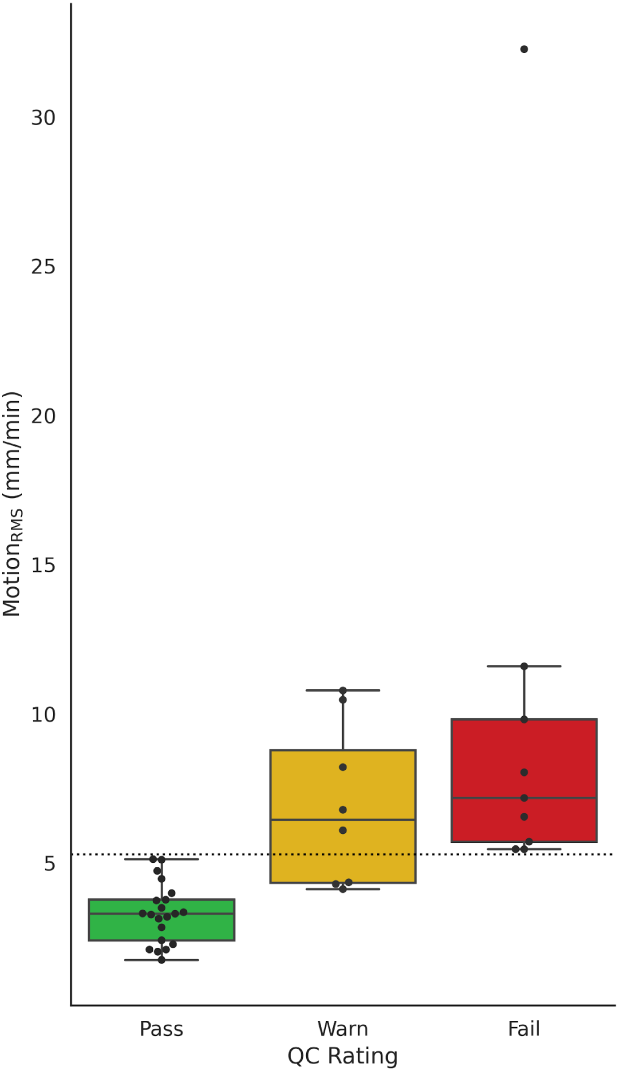
Head motion during structural MRI as a function of manual image quality ratings. A boundary was established between the upper end of pass ratings and the lower end of fail ratings (Motion_RMS_ = 5.29 mm/min).

As shown in Figure 7, the head mold improved scan quality for many participants, with five remaining poor or getting worse. The scans of six participants (2 TDC, 4 with ADHD) failed QC without the head mold but passed QC with the head mold (fail - pass). There were four individuals (3 TDC, 1 with ADHD) who had unusable data for both conditions (fail - fail). There were eight individuals (7 TDC, 1 with ADHD) whose data quality was good whether or not they used the head mold (pass - pass). Finally, one TDC participant (9 years old, low anxiety) passed QC without the head mold but failed QC with the head mold on (pass - fail). This participant was scanned without the head mold first, and they reported that both scan conditions were equally tolerable (happy face on the Wong-Baker scale) and that the MOLD+ condition was preferred over MOLD-condition.

**Fig. 7.**
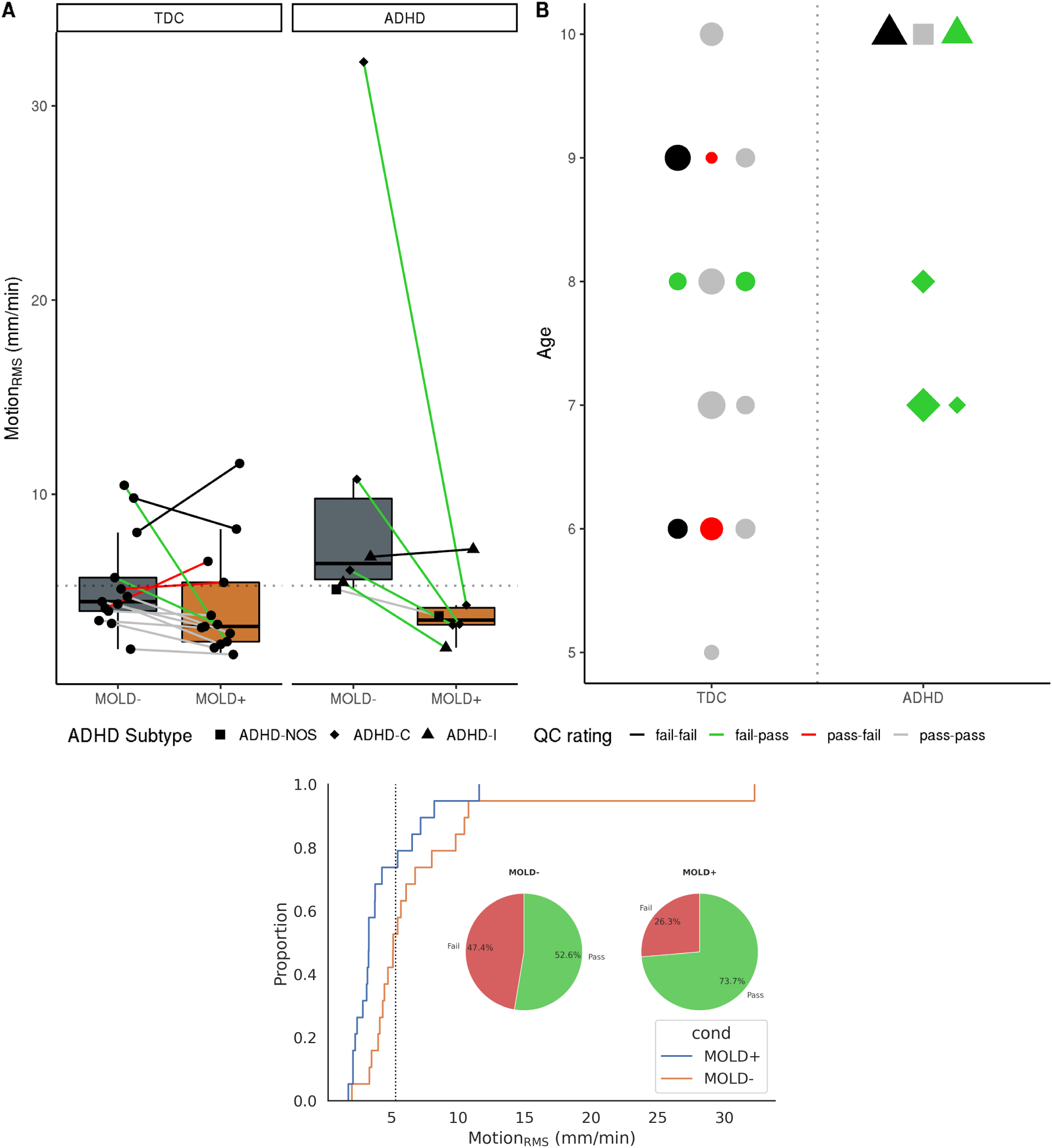
Characterizing head motion during sMRI scan and its impact on data quality. **A.** Mean motion (Motion_RMS_) with and without the head mold. Each colored line resembles within-participant change between MOLD- and MOLD+. The colors of the lines indicate how the data quality was affected by the head mold at a given QC rating threshold. The horizontal dotted line denotes the QC threshold used for defining data quality (Motion_RMS_ < 5.29 mm/min). **B.** Head mold-related impacts on data quality as a function of age, ADHD diagnosis, and anxiety. Each marker resembles individual participants, and the colors denote the changes in QC ratings between MOLD- and MOLD+. The size of the symbols denotes the total anxiety score from SCARED. **C.** A cumulative distribution function showing that MOLD+ scans have a higher probability of having a lower mean FD compared to MOLD-scans. At Motion_RMS_ = 5.29 mm/min (vertical dashed line in black), the head mold increased the percentage of usable scans from 52.6% to 74%.

As in the case with FD during functional scans, we observed a higher proportion of structural scans with lower Motion_RMS_ during the MOLD+ condition as compared to the MOLD-condition as indicated in Figure 7C. At the QC threshold of Motion_RMS_ = 5.29 mm/min, the head mold increased the percentage of total usable scans from 52.6% to 74%.

Without the head mold (MOLD-), children with ADHD had higher head motion than TDC though it did not reach statistical significance (*t*(18) = 1.89, *p* = 0.075). Motion appeared higher for the ADHD-C subtype (N = 3) compared to the ADHD-I subtype (N = 2), though there were too few observations for statistical testing. Age and total anxiety were not significantly predictive of motion (r = −0.14; r = −0.14, respectively). With the head mold on (MOLD+), there were no observable differences in head motion between ADHD and TDC groups (*t*(18) = 0.29, *p* = 0.78). The head mold reduced head motion and improved image quality in 4 of the 6 children with ADHD. Of the remaining two, one child had good image quality in both MOLD+ and MOLD-, and the other had poor quality data in both conditions. Neither total anxiety score nor age were associated with the success of the head mold for improving data quality (r = 0.12 for both).

Manual ratings based on qualitative assessments are currently standard practice for assessing the quality of sMRI scans; however, a number of measures have been proposed to quantitatively assess image quality. We evaluated the impact of the head mold on measures of image quality generated by CAT12 and FreeSurfer’s Euler number (Figure 8). The Euler number is derived from the number of holes in the reconstructed cortical surface (Dale et al., 1999). Therefore, a lower Euler number indicates a higher image quality (fewer cortical holes), and visa versa. The Euler number has been reported to be a reliable measure of data quality, and it has been validated against other tools and with manual ratings (Rosen et al., 2018). The CAT12 image quality rating (IQR) combines various parameters of noise and spatial resolution obtained during its preprocessing procedures. Although IQR appeared to increase with the head mold on, it did not meet statistical significance (F_(1, 18)_ = 2.84, p > 0.05). Likewise, there was no significant difference in Euler number between the scan conditions (F_(1, 18)_ = 2.44, p > 0.05), despite a general decreasing trend.

**Fig. 8.**
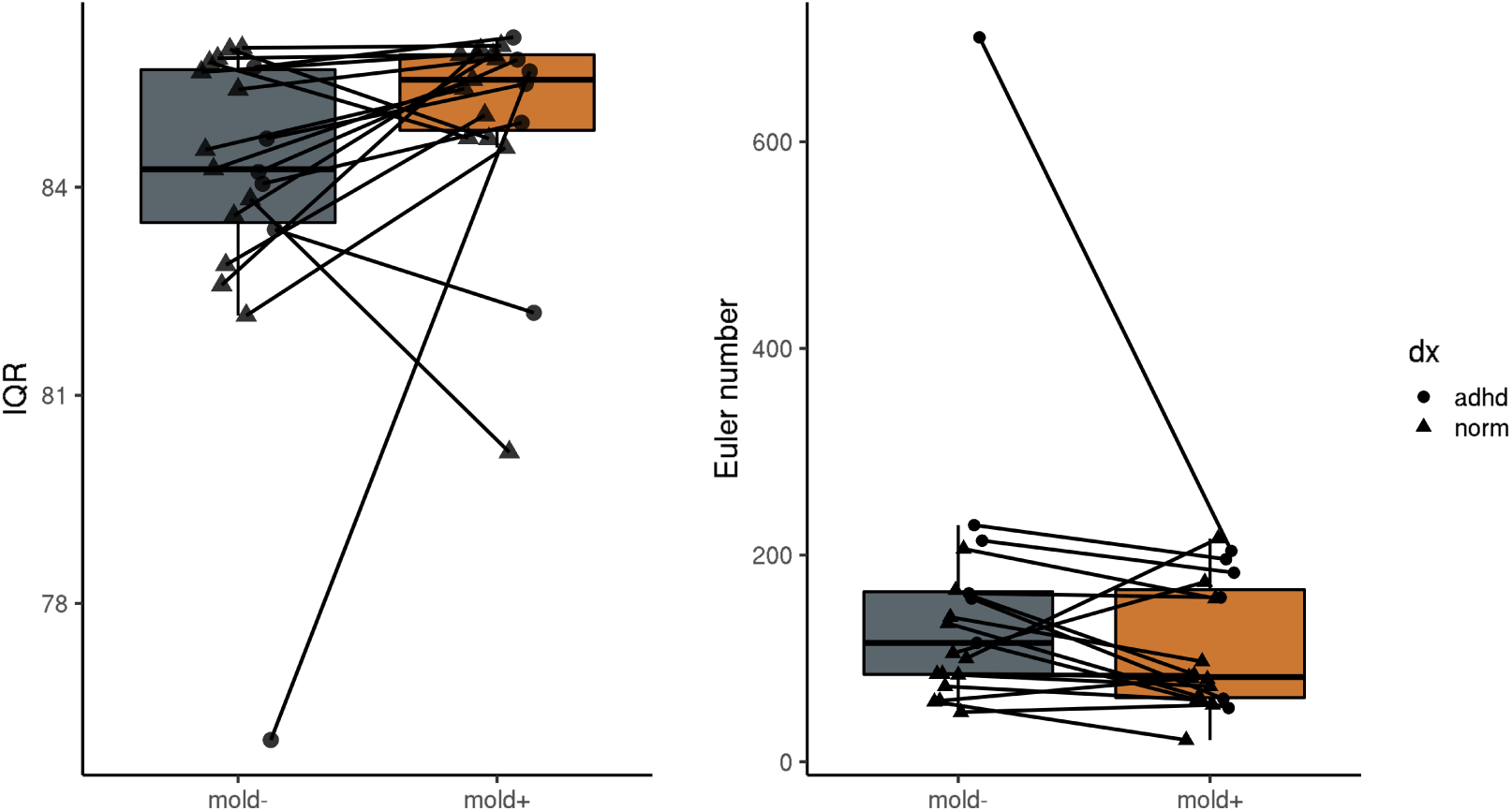
Assessing the impact of head molds on quantitative estimates of image quality. Image Quality Rating (IQR) is a composite measure from CAT12 and the Euler number is Freesurfer’s estimates of cortical holes.

#### 3.4.3 Session Effects

Finally, we evaluated the effect of scan order on the head mold performance (Figure 9). We found that the scan order did not affect head motion for the MOLD-condition for either the sMRI or the fMRI scan (*p’s* > 0.05). Participants who wore the head mold during the second scan tended to move more than those who had the head mold on during the first scan; however, the results did not reach statistical significance (*p’s* > 0.05).

**Fig. 9.**
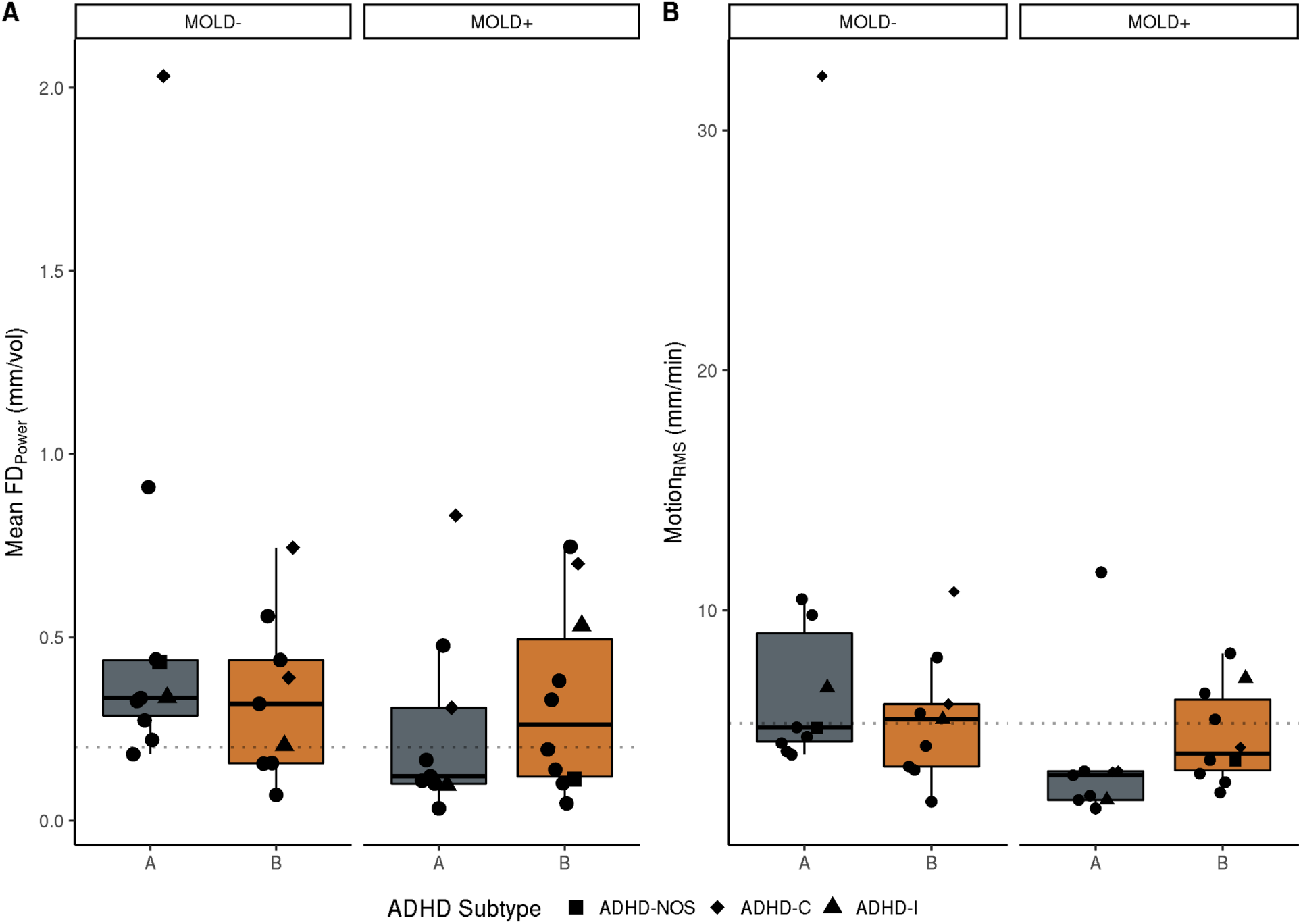
Examining the effect of scan order for **A.** functional MRI scans and **B.** structural MRI scans. For each scan type the scan order effects are displayed for MOLD- and MOLD+ conditions separately.

### 3.5 How do head molds impact analysis results?

#### 3.5.1 Functional

Distance-dependent effects of motion on functional connectivity estimates have been well established (Power et al., 2012; Satterthwaite et al., 2012). Functional connectivity (FC) between short-distance ROI pairs tends to be artifactually increased by motion and FC between long-distance ROI pairs tends to be reduced (Power et al., 2012). To this end, we examined whether the head mold would impact the distance-dependent relationship between head motion and signal covariance. We assessed the relationship between distance and time series correlations for the two scan conditions, averaged across individuals (Fig 10). Both scan conditions showed the expected relationship between FC and distance where local correlations are higher than correlations between distant regions. The head mold did not globally ameliorate the negative impacts of motion on FC across the sample. However, when we examined individual participant data, we observed that the distance-FC relationship was improved in participants whose head motion was reduced by the head mold, particularly in cases where the QC ratings were converted from fail to pass with the head mold on (Fig 10B, middle row).

**Fig. 10.**
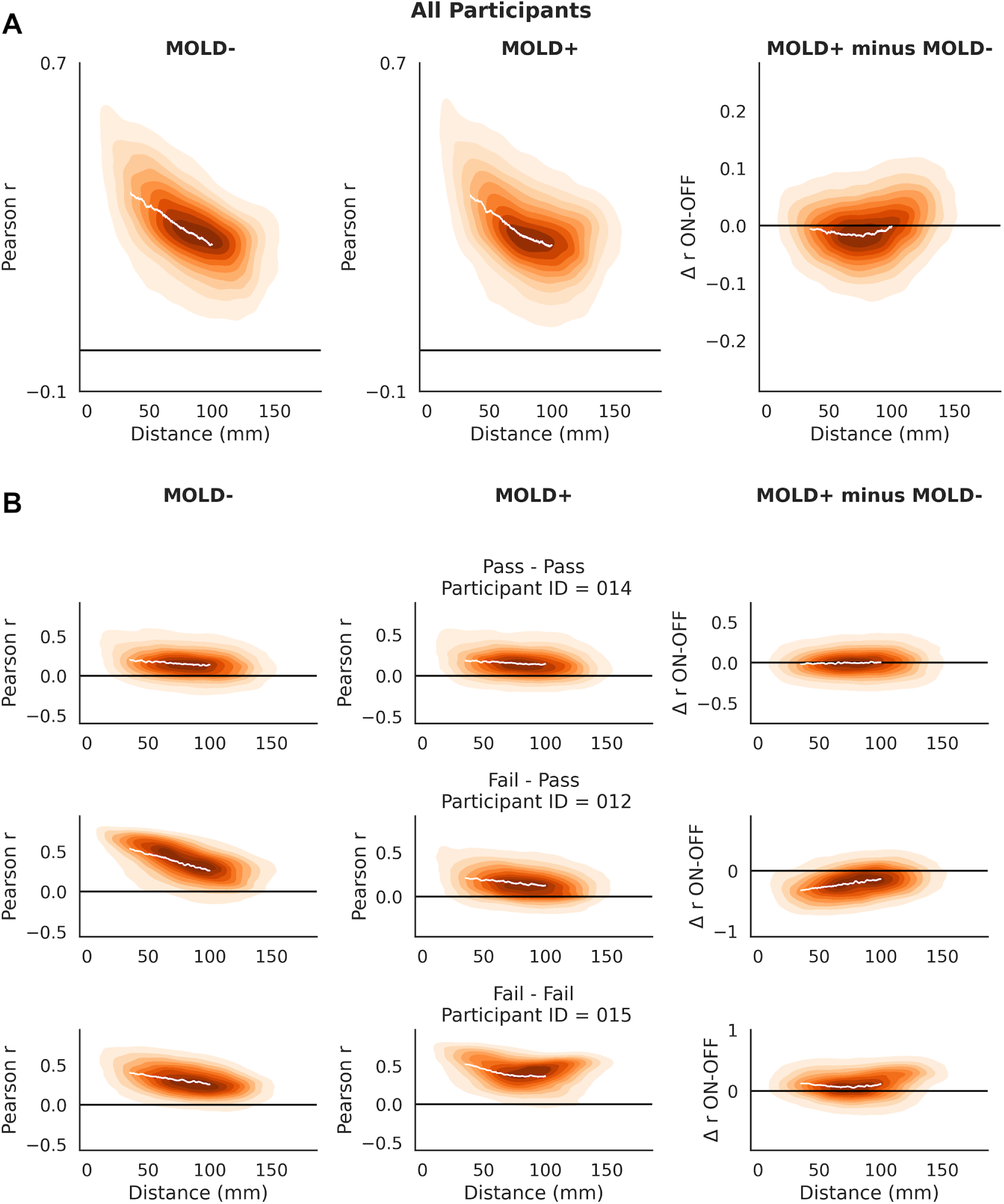
**A.** Distance-dependent artifacts in functional connectivity analyses. Group-averaged plots depicting that head motion inflates correlations of nearby regions more than distant regions. The plot on the right panel shows that the distance-related artifact levels off toward more distant regions. These plots show the relationship between distance and correlation values and how they differ between MOLD+ and MOLD-conditions. The difference plot on the bottom right indicates little change in correlation values between the head mold conditions. **B.** Participant-level patterns of distance-FC relationship. Note: The scans shown in this figure correspond with the motion traces from Figure 4. These are all TDC individuals with low anxiety scores.

#### 3.5.2 Structural

Percent brain volume was significantly lower in the MOLD+ condition compared to the MOLD-condition (Figure 11A; *t*(18) = −8.00, *p* < 0.000001). Linear mixed effects (LME) models were fitted to the data in order to test for an interaction between head mold condition and head motion on gray matter volume estimates. For both CAT12 and FreeSurfer, significant interactions between head mold condition and head motion were observed (CAT12: *F*_(1,16)_ = 4.78, *p* < 0.05; FreeSurfer *F*_(1,16)_ = 10.45, *p* < 0.01; Figure 11B and 11C). For both algorithms, we observed a negative association between head motion and GM estimates which was stronger for the MOLD-condition compared to MOLD+.

**Fig 11.**
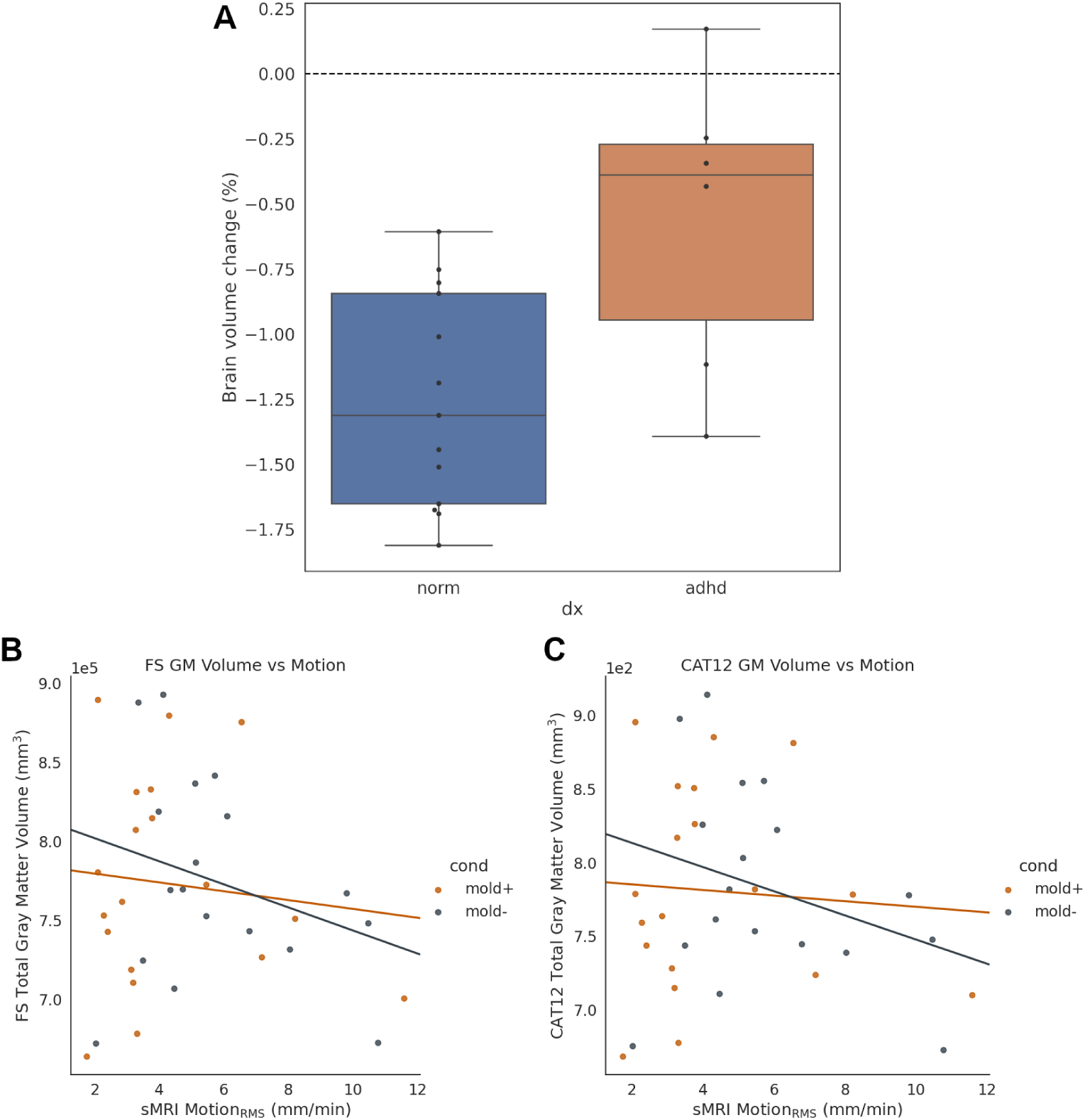
**A.** Percent brain volume change (as estimated by FSL Siena) after the head mold was placed on. **B and C**. Relationship between gray matter volume estimates and head motion (RMS displacement) as a function of the head mold condition (MOLD- vs MOLD+). Both imaging softwares (**B:** Freesurfer; **C**: CAT12) exhibited a similar pattern of results.

## 4. Discussion

Head motion during MRI scanning remains a persistent challenge to scientific findings, particularly in hyperkinetic populations such as young children with or without neurodevelopmental disorder. In the present study, we build on previous studies by testing the efficacy of using a custom-fitted head mold to reduce head motion during fMRI and sMRI acquisition in 19 young children (ages 5 - 10), six of whom had ADHD. Following Horien et al. (2020), we utilized a combination of techniques to maximize head motion reductions in pediatric populations, including mock scanning with thorough training, film watching during scanning, and using a weighted blanket. We assessed the impact of the head mold on head motion, data quality, and analysis results derived from the scans. Indeed, consistent with previous literature, the children in our sample exhibited high motion; 15 out of 19 fMRI scans (78.9%) and 10 out of 19 sMRI scans (52.6%) were contaminated with suprathreshold motion that would be excluded based on conventional QC approaches. We found that the head mold benefits were not universal in our sample; some benefited while others did not, and one sMRI scan was worsened by the mold. Whether or not a head mold would mitigate motion, or improve data quality, for a participant could not be predicted by age, ADHD diagnosis, or anxiety; however the sample size is likely too small to adequately evaluate this type of prediction. The head molds were well tolerated by the participants, and they provided ample hearing protection when used in combination with Sensimetrics ear monitors.

Custom-fitted head molds were first reported by Power et al. (2019) to reduce head motion in resting-state fMRI data in typically-developed participants ranging from 7 - 28 years of age (mean age = 15 years). In a later study, Jolly et al. (2020) demonstrated a more limited utility of the head molds for reducing head motion during a naturalistic viewing task and a spoken recall task in young adults (mean age = 23 years). We extend this previous work by investigating whether head molds can improve head motion and data quality specifically in younger children (5 - 10 years old), including six with ADHD. Neurodevelopment and neurodevelopmental disorders are two areas that are likely to benefit substantially from neuroimaging research, yet they represent the hardest populations for conducting this type of research due to head motion (Alexander-Bloch et al., 2016; Pardoe et al., 2016; Satterthwaite et al., 2012). Therefore, reducing head motion in these populations is highly valuable.

Our results show that head mold-related reductions in head motion were not universal; we found considerable variability unlike Power et al. (2019) where decreases in head motion were found in nearly all of their participants. In the present study, we found evidence that head mold performance could change over the course of a scanning session, putatively due to fatigue. Participants who wore the head mold as their second scan tended to move more than those who wore the mold during the first scan. Although the results were not significantly significant, it suggests that head mold performance may be optimal for shorter scan sessions. Further, this effect could contribute to variability in head mold performance as a function of study design. For example, a multi-session or cross-sectional evaluation of head mold effects may receive different results.

Another possible contribution to the high variability in head mold performance in the present study could be related to the control condition consisting of more comprehensive motion reduction techniques than a typical scan. As such, this could create a floor effect that may account for motion reductions that were not as drastically apparent as those found by Power et al. (2019). Paired t-tests between the head motion from scans acquired with and without head mold, across all participants, approached significance for both fMRI (*p* = 0.08) and sMRI (*p* = 0.07), but change in head motion between scan conditions may not provide complete insight into the head mold effects.

Therefore, rather than focusing exclusively on reductions in motion, we also assessed whether the head mold would increase the number of scans that would pass conventional QC procedures. This approach better highlights the benefits of using the head mold towards data quality. For example, we observed cases where motion was reduced by the head mold but the scans would have passed QC with or without the head mold (pass-pass). In these scenarios, the overall effect of the head mold could be minimal. On the other hand, the situations that were most valuable were when scans failed QC during MOLD- but passed QC during MOLD+ (fail-pass). Both sMRI and fMRI scans benefited in this way (N = 6 and N = 7, respectively); however, the fail-pass individuals were not exactly the same between the scan modalities. For fMRI scans, we observed that image quality benefited more in the TDC group than the ADHD group, presumably due to higher motion overall in the ADHD group.

The QC threshold for fMRI data was based on mean FD, and the threshold for sMRI data was based on a 5-point manual rating system that was then classified as Pass or Fail. Using measurements from the vNAV sequence, we verified that sMRI image quality was closely related to head motion during sMRI scanning, rather than other artifacts. This suggests that head motion translates well into image quality, and it further supports the importance of reducing head motion to improve data quality.

In addition to image quality, we evaluated the impact of the head molds on MRI-based estimates commonly employed in neuroimaging analyses. For the fMRI scans, we examined FC between pairs of ROIs and whether the head mold reduced the impact of distance between ROI pairs on FC. The head mold did not ameliorate the distance-FC relationship in all cases; however we observed that it reduced the impact of distance on FC when it reduced head motion. This result differed from Power et al. (2019) who reported an average reduction in the distance-dependent artifact across their entire sample.

For the sMRI scans, we assessed the impact of the head mold on metrics of brain morphometry. Reuter et al. (2015) reported that head motion negatively correlated with brain morphometry metrics. Therefore, we tested the hypothesis that the head mold would reduce sMRI head motion and translate to higher morphometry metrics compared to the control condition. Although the head mold reduced sMRI head motion for the majority of the participants, we observed the head mold decreased percent brain volume estimates as performed by FSL Siena, a result that was not consistent with the findings of Reuter et al. (2015). A similar pattern of findings emerged for the gray matter volume estimates by FreeSurfer and SPM12 CAT. Our results suggest that the morphometry metrics were overestimated due to head motion, rather than underestimated as was reported by Reuter et al. (2015).

We also assessed the extent to which head motion was correlated between fMRI and sMRI scanning. Savalia et al. (2017) previously reported that head motion during fMRI can reliably predict the quality of sMRI scans from the same session. Based on their results, the authors proposed that fMRI motion could serve as a proxy for sMRI motion since it cannot be measured with conventional sMRI sequences. We directly tested this hypothesis using vNAV-derived head motion during sMRI scanning. Indeed, without the head mold on, there was a high linear correlation between fMRI motion and sMRI motion. However, despite the high correlation, the motion was not equivalent between scans based on a low concordance correlation coefficient (CCC). With the head mold on, the motion was also correlated, although not as highly as without the mold, and the head mold increased CCC between the scans. This result suggests that the head mold equalized head motion between scans more so than without the head mold.

Due to the nature of how the head mold fit around participants’ heads and within the scanning coil, we were unable to use our typical hearing protection strategy. Using acoustical measurements, we demonstrated that Caseforge head molds amply protected participants’ hearing when used in combination with earplugs. Indeed, an early study by Ravicz and Melcher (2001) advocated the idea of using a helmet, in combination with earplugs and earmuffs, as a tool for additional hearing protection. In their study, the helmet was not designed to restrict head motion, rather it served as an added barrier between the scanner noises and the participants’ head presumably reducing sound conduction along both the head and ear canal routes. In agreement with Ravicz and Melcher (2001), we found that the combination of head mold, earplug, and foam padding provided superior reduction in noise dosage compared to earplugs alone. The head mold used in the present study bolstered hearing protection while also reducing head motion despite only being designed for immobilization purposes.

### Practical Considerations and Future Directions

In carrying out the present study, several issues arose that are worth noting. The Styrofoam head mold frequently needed to be modified extemporaneously in between scan conditions because they did not fit properly. Fortunately, the Styrofoam material allowed for quick modifications using a utility knife to shave down problematic areas. This occurred in almost half of our sample (9 out of 19, ∼47%). We have reasoned that this could be due to minor errors in the 3D scanning of participants’ heads, errors in the 3D printing of the head mold, or from rapid head growth during the time elapsed between when the molds were created and when participants were scanned (mean time elapsed = 18.6 ± 10.3 days). Due to time constraints in the present study, we did not take advantage of the iterative and interactive fitting process that the Caseforge company offers.

Participants in the present study completed a 60-minute scanning protocol, during which the head mold was on for 30 minutes. The duration limits of the molds that are tolerated by participants remains unknown and would be useful to test in future studies. Another outstanding issue pertains to identifying factors that predict who might benefit from the mold and who might not. In the present study, motion could not be reliably predicted by age, ADHD status, or parent-reported ratings of anxiety. Further research is needed in order to determine which populations, task conditions, etc., might benefit the most from using the head mold. Finally, the efficacy of the head mold on task-based fMRI using conventional cognitive neuroscience paradigms remains to be determined. To date, existing studies using the head mold (including the present study) include film watching (Jolly et al., 2020), resting-state cross-hair fixation (Power et al., 2019), and a spoken recall task (Jolly et al., 2020). More studies will be required to determine how the head molds perform across a variety of tasks.

## Conclusions

Taken together, the head molds used in the present study offer an encouraging method for reducing head motion in the developmental population, including those with neurodevelopmental disorders. The custom-fitted head mold may not be a universal solution for reducing head motion-related artifacts, but we believe the additional time and cost (∼$125) required for the head mold outweighs the potential loss of data from excessive head motion. The comprehensive fitting process using 3D optical scanning ensured that the mold fit comfortably around participants’ heads. Indeed, the head mold was well-tolerated by our sample of young participants, some of whom had ADHD. A large majority of participants indicated their willingness to return for another scan using the head mold. Determining predictors of who will benefit from the head mold will be valuable for future studies across a diversity of study samples, tasks, and scanning conditions.

## Data Availability

The datasets generated during and/or analysed during the current study are available from the corresponding author on reasonable request.

## Acknowledgements

This work was supported by startup funding to Cameron Craddock.

## Supplemental Information

The Supplemental Information contains details and results of the acoustic measurements taken for this study. Figs. S1 and S2 depict the experimental setup, Fig. S3 shows the results comparing the effectiveness of the various combinations of hearing protection, and Fig. S4 shows the sound levels a participant would be exposed to during an sMRI and fMRI scan when using selected combinations of hearing protection. These results were then applied to sMRI and fMRI scans of different lengths to determine the resulting noise dose (Table S1), which can be compared to established OSHA regulations for exposure to sound, to understand the effectiveness of hearing protection in the context of typical scanning routines.

**Fig. S1.**
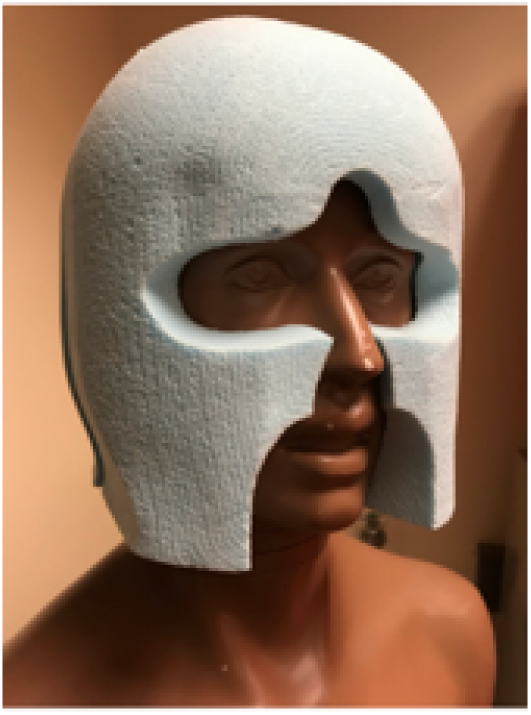
Caseforge head mold on KEMAR acoustic research manikin.

**Fig. S2.**
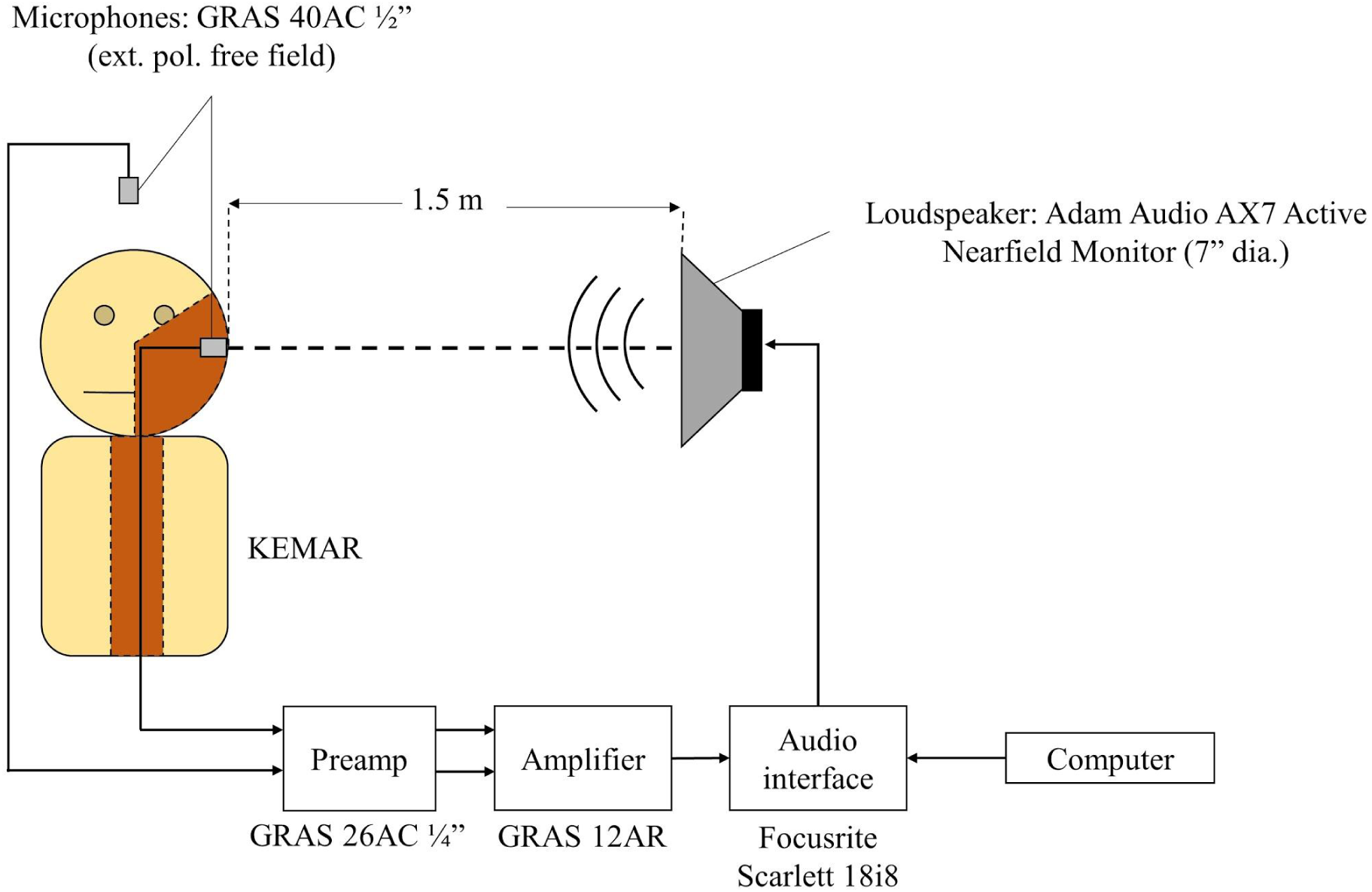
Experimental setup for hearing protection evaluation.

**Fig. S3.**
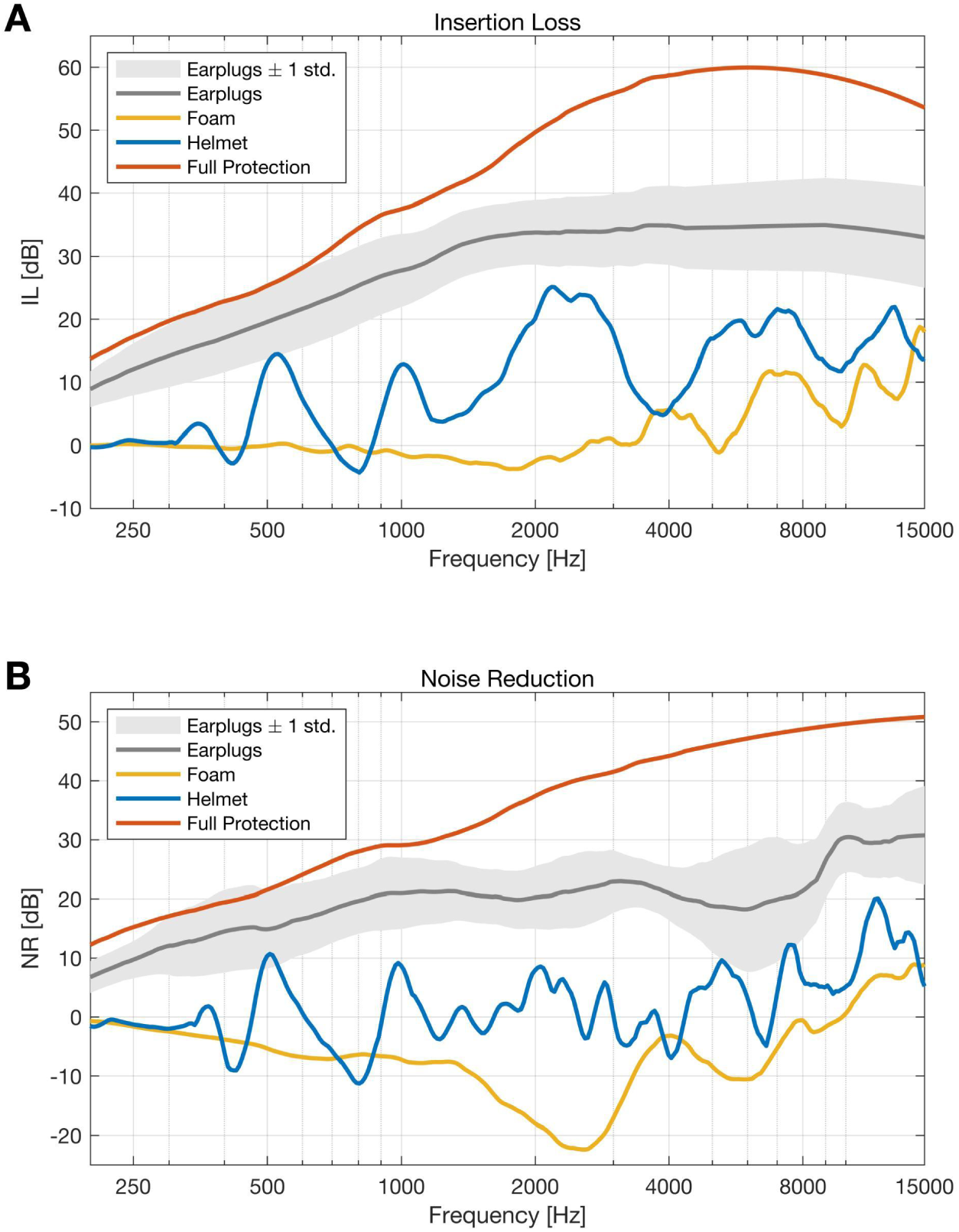
**A.** Insertion loss (IL) properties for the various combinations of hearing protection devices. One standard deviation range is shown for the earplugs to account for the fact that the earplugs may be inserted differently depending on the patient. **B.** Noise reduction (NR) properties for the various combinations of hearing protection devices. Note that NR contains the effects of the KEMAR head-related transfer function.

**Fig. S4.**
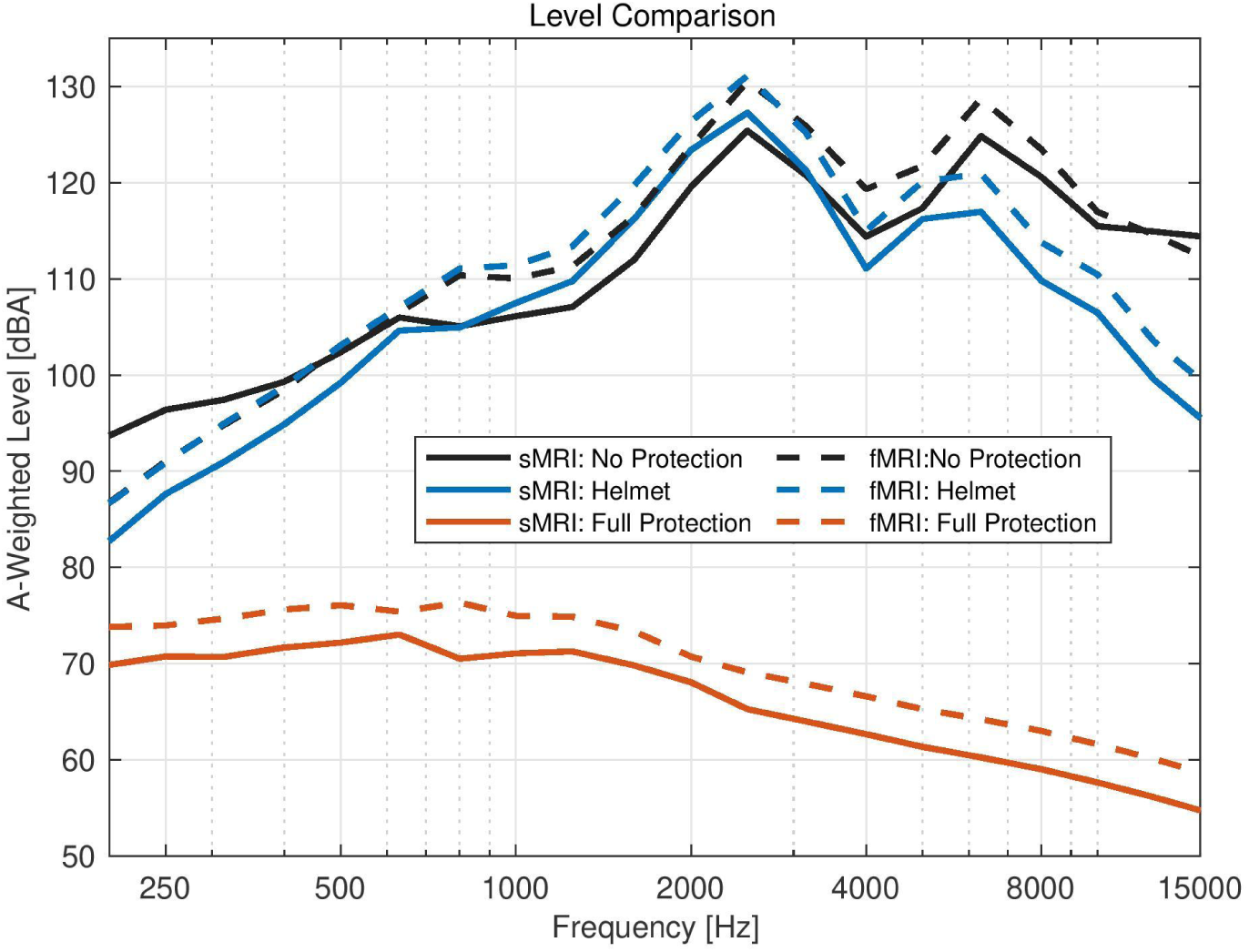
Human head-related transfer function applied to KEMAR shows estimated frequency content that KEMAR would receive inside the MRI machine while scanning. Levels (dB re 20 µPa) are in 1/3 octave bands with center frequencies and A-weighting provided by Long (2014).

**Table S1.** Noise dosage with earplugs and head mold over varying durations of scanning. The Criterion Level and Exchange Level used in these calculations are 90 dB and 5 dB, respectively. Dose is given as a percentage of the total allowable exposure to sound at a given SPL in an 8-hour period. A value of 100% indicates the maximum allowable time of exposure at a given SPL. *Configuration used in the present study.

|  |  |  | fMRI |  |  |
| --- | --- | --- | --- | --- | --- |
|  | None | Earphones Only | Head Mold Only | Earphones + Head Mold | Earphones + Head Mold + Foam Padding* |
| **SPL RMS**<br>[re 20 μPa] | 115.18 dB | 81.58 dB | 108.22 dB | 79.82 dB | 73.19 dB |
| **15 min** | 102.55 | 0.97% | 39.08% | 0.76% | 0.30% |
| **30 min** | 205.11% | 1.94% | 78.16% | 1.52% | 0.61% |
| **45 min** | 307.66% | 2.92% | 117.24% | 2.28% | 0.91% |
| **60 min** | 410.21 | 3.89% | 156.32% | 3.05% | 1.21% |
|  |  |  | sMRI |  |  |
|  | None | Earphones Only | Head Mold Only | Earphones + Head Mold | Earphones + Head Mold + Foam Padding* |
| **SPL RMS**<br>[re 20 μPa] | 114.80 dB | 80.67 dB | 100.55 dB | 76.14 dB | 70.80 dB |
| **2.5 min** | 16.21% | 0.14% | 2.25% | 0.08% | 0.04% |
| **5 min** | 32.41% | 0.29% | 4.50% | 0.15% | 0.07% |
| **7.5 min** | 48.62% | 0.43% | 6.75% | 0.23% | 0.11% |
| **10 min** | 64.82% | 0.57% | 9.00% | 0.30% | 0.15% |

## Footnotes

1 Mean FD < 0.2 mm has been proposed as criteria for assessing the overall quality of an entire functional scan (Ciric et al., 2017), whereas FD < 0.3 mm was used as a criteria for identifying individual motion spikes for subsequent spike regression or scrubbing. The 0.3 mm scrubbing threshold was chosen to resemble Power et al. (2019).

## References

Alexander, L. M., Escalera, J., Ai, L., Andreotti, C., Febre, K., Mangone, A., Vega-Potler, N., Langer, N., Alexander, A., Kovacs, M., Litke, S., O’Hagan, B., Andersen, J., Bronstein, B., Bui, A., Bushey, M., Butler, H., Castagna, V., Camacho, N., … Milham, M. P. (2017). An open resource for transdiagnostic research in pediatric mental health and learning disorders. Scientific Data, 4(1), 170181. https://doi.org/10.1038/sdata.2017.181

Alexander-Bloch, A., Clasen, L., Stockman, M., Ronan, L., Lalonde, F., Giedd, J., & Raznahan, A. (2016). Subtle in-scanner motion biases automated measurement of brain anatomy from in vivo MRI. Human Brain Mapping, 37(7), 2385–2397. https://doi.org/10.1002/hbm.23180

Badachhape, A. A., Okamoto, R. J., Johnson, C. L., & Bayly, P. V. (2018). Relationships between scalp, brain, and skull motion estimated using magnetic resonance elastography. Journal of Biomechanics, 73, 40–49. https://doi.org/10.1016/j.jbiomech.2018.03.028

Benner, T., van der Kouwe, A. J. W., & Sorensen, A. G. (2011). Diffusion imaging with prospective motion correction and reacquisition. Magnetic Resonance in Medicine, 66(1), 154–167. https://doi.org/10.1002/mrm.22837

Blumenthal, J. D., Zijdenbos, A., Molloy, E., & Giedd, J. N. (2002). Motion Artifact in Magnetic Resonance Imaging: Implications for Automated Analysis. NeuroImage, 16(1), 89–92. https://doi.org/10.1006/nimg.2002.1076

Caballero-Gaudes, C., & Reynolds, R. C. (2017). Methods for cleaning the BOLD fMRI signal. NeuroImage, 154, 128–149. https://doi.org/10.1016/j.neuroimage.2016.12.018

Ciric, R., Wolf, D. H., Power, J. D., Roalf, D. R., Baum, G. L., Ruparel, K., Shinohara, R. T., Elliott, M. A., Eickhoff, S. B., Davatzikos, C., Gur, R. C., Gur, R. E., Bassett, D. S., & Satterthwaite, T. D. (2017). Benchmarking of participant-level confound regression strategies for the control of motion artifact in studies of functional connectivity. NeuroImage, 154, 174–187. https://doi.org/10.1016/j.neuroimage.2017.03.020

Couvy-Duchesne, B., Ebejer, J. L., Gillespie, N. A., Duffy, D. L., Hickie, I. B., Thompson, P. M., Martin, N. G., Zubicaray, G. I. de, McMahon, K. L., Medland, S. E., & Wright, M. J. (2016). Head Motion and Inattention/Hyperactivity Share Common Genetic Influences: Implications for fMRI Studies of ADHD. PLOS ONE, 11(1), e0146271. https://doi.org/10.1371/journal.pone.0146271

Cox, R. W. (1996). AFNI: software for analysis and visualization of functional magnetic resonance neuroimages. Computers and Biomedical Research, 29, 162–173.

Cox, R. W., & Jesmanowicz, A. (1999). Real-time 3D image registration for functional MRI. Magnetic Resonance in Medicine, 42(6), 1014–1018. https://doi.org/10.1002/(SICI)1522-2594(199912)42:6<1014::AID-MRM4>3.0.CO;2-F

Dale, A. M., Fischl, B., & Sereno, M. I. (1999). Cortical surface-based analysis. I. Segmentation and surface reconstruction. NeuroImage, 9(2), 179–194. https://doi.org/10.1006/nimg.1998.0395

Davis, M. A. (2016). Aromatherapy in MRI - Evaluating the use of Elequil Aromatabs to Reduce Aborted Scans in Patients with Anxiety and Claustrophobia (p. 4).

de Bie, H. M. A., Boersma, M., Wattjes, M. P., Adriaanse, S., Vermeulen, R. J., Oostrom, K. J., Huisman, J., Veltman, D. J., & Delemarre-Van de Waal, H. A. (2010). Preparing children with a mock scanner training protocol results in high quality structural and functional MRI scans. European Journal of Pediatrics, 169(9), 1079–1085. https://doi.org/10.1007/s00431-010-1181-z

Dosenbach, N. U. F., Koller, J. M., Earl, E. A., Miranda-Dominguez, O., Klein, R. L., Van, A. N., Snyder, A. Z., Nagel, B. J., Nigg, J. T., Nguyen, A. L., Wesevich, V., Greene, D. J., & Fair, D. A. (2017). Real-time motion analytics during brain MRI improve data quality and reduce costs. NeuroImage, 161, 80–93. https://doi.org/10.1016/j.neuroimage.2017.08.025

Ducharme, S., Albaugh, M. D., Nguyen, T.-V., Hudziak, J. J., Mateos-Pérez, J. M., Labbe, A., Evans, A. C., & Karama, S. (2016). Trajectories of cortical thickness maturation in normal brain development – The importance of quality control procedures. NeuroImage, 125, 267–279. https://doi.org/10.1016/j.neuroimage.2015.10.010

Engelhardt, L. E., Roe, M. A., Juranek, J., DeMaster, D., Harden, K. P., Tucker-Drob, E. M., & Church, J. A. (2017). Children’s head motion during fMRI tasks is heritable and stable over time. Developmental Cognitive Neuroscience, 25, 58–68. https://doi.org/10.1016/j.dcn.2017.01.011

Epstein, J. N., Casey, B. J., Tonev, S. T., Davidson, M., Reiss, A. L., Garrett, A., Hinshaw, S. P., Greenhill, L. L., Vitolo, A., Kotler, L. A., Jarrett, M. A., & Spicer, J. (2007). Assessment and prevention of head motion during imaging of patients with attention deficit hyperactivity disorder. Psychiatry Research: Neuroimaging, 155(1), 75–82. https://doi.org/10.1016/j.pscychresns.2006.12.009

Esper, N., Much, M., Azevedo, D., Buchweitz, A., Milham, M., & Franco, A. R. (2019, June). Real-time fMRI Motion Tracking: Should I stop and restart the scan? [Poster Presentation]. OHBM.

Friston, K. J., Williams, S., Howard, R., Frackowiak, R. S. J., & Turner, R. (1996). Movement-Related effects in fMRI time-series. Magnetic Resonance in Medicine, 35(3), 346–355. https://doi.org/10.1002/mrm.1910350312

Gaser, C., & Dahnke, R. (2016). CAT - A Computational Anatomy Toolbox for the Analysis of Structural MRI Data. Organization of Human Brain Mapping. https://www.neuro.uni-jena.de/hbm2016/GaserHBM2016.pdf

Godenschweger, F., Kägebein, U., Stucht, D., Yarach, U., Sciarra, A., Yakupov, R., Lüsebrink, F., Schulze, P., & Speck, O. (2016). Motion correction in MRI of the brain. Physics in Medicine and Biology, 61(5), R32–R56. https://doi.org/10.1088/0031-9155/61/5/R32

Greene, D. J., Koller, J. M., Hampton, J. M., Wesevich, V., Van, A. N., Nguyen, A. L., Hoyt, C. R., McIntyre, L., Earl, E. A., Klein, R. L., Shimony, J. S., Petersen, S. E., Schlaggar, B. L., Fair, D. A., & Dosenbach, N. U. F. (2018). Behavioral interventions for reducing head motion during MRI scans in children. NeuroImage, 171, 234–245. https://doi.org/10.1016/j.neuroimage.2018.01.023

Harris, P. A., Taylor, R., Minor, B. L., Elliott, V., Fernandez, M., O’Neal, L., McLeod, L., Delacqua, G., Delacqua, F., Kirby, J., & Duda, S. N. (2019). The REDCap consortium: Building an international community of software platform partners. Journal of Biomedical Informatics, 95, 103208. https://doi.org/10.1016/j.jbi.2019.103208

Harris, P. A., Taylor, R., Thielke, R., Payne, J., Gonzalez, N., & Conde, J. G. (2009). Research electronic data capture (REDCap)—A metadata-driven methodology and workflow process for providing translational research informatics support. Journal of Biomedical Informatics, 42(2), 377–381. https://doi.org/10.1016/j.jbi.2008.08.010

Holdsworth, S. J., Aksoy, M., Newbould, R. D., Yeom, K., Van, A. T., Ooi, M. B., Barnes, P. D., Bammer, R., & Skare, S. (2012). Diffusion tensor imaging (DTI) with retrospective motion correction for large-scale pediatric imaging. Journal of Magnetic Resonance Imaging, 36(4), 961–971. https://doi.org/10.1002/jmri.23710

Horien, C., Fontenelle, S., Joseph, K., Powell, N., Nutor, C., Fortes, D., Butler, M., Powell, K., Macris, D., Lee, K., Greene, A. S., McPartland, J. C., Volkmar, F. R., Scheinost, D., Chawarska, K., & Constable, R. T. (2020). Low-motion fMRI data can be obtained in pediatric participants undergoing a 60-minute scan protocol. Scientific Reports, 10(1), 21855. https://doi.org/10.1038/s41598-020-78885-z

Hu, Y., & Glover, G. H. (2007). Three-dimensional spiral technique for high-resolution functional MRI. Magnetic Resonance in Medicine, 58(5), 947–951. https://doi.org/10.1002/mrm.21328

Jolly, E., Sadhukha, S., & Chang, L. J. (2020). Custom-molded headcases have limited efficacy in reducing head motion during naturalistic fMRI experiments. NeuroImage, 222, 117207. https://doi.org/10.1016/j.neuroimage.2020.117207

Kreilkamp, B. A. K., Weber, B., Richardson, M. P., & Keller, S. S. (2017). Automated tractography in patients with temporal lobe epilepsy using TRActs Constrained by UnderLying Anatomy (TRACULA). NeuroImage: Clinical, 14, 67–76. https://doi.org/10.1016/j.nicl.2017.01.003

Le Bihan, D., Poupon, C., Amadon, A., & Lethimonnier, F. (2006). Artifacts and pitfalls in diffusion MRI. Journal of Magnetic Resonance Imaging, 24(3), 478–488. https://doi.org/10.1002/jmri.20683

Liu, G., Sobering, G., Duyn, J., & Moonen, C. T. W. (1993). A functional MRI technique combining principles of echo-shifting with a train of observations (PRESTO). Magnetic Resonance in Medicine, 30(6), 764–768. https://doi.org/10.1002/mrm.1910300617

Long, M. (2014). Architectural Acoustics. Elsevier. https://doi.org/10.1016/C2009-0-64452-4

Maclaren, J., Armstrong, B. S. R., Barrows, R. T., Danishad, K. A., Ernst, T., Foster, C. L., Gumus, K., Herbst, M., Kadashevich, I. Y., Kusik, T. P., Li, Q., Lovell-Smith, C., Prieto, T., Schulze, P., Speck, O., Stucht, D., & Zaitsev, M. (2012). Measurement and Correction of Microscopic Head Motion during Magnetic Resonance Imaging of the Brain. PLOS ONE, 7(11), e48088. https://doi.org/10.1371/journal.pone.0048088

Maclaren, J., Herbst, M., Speck, O., & Zaitsev, M. (2013). Prospective motion correction in brain imaging: A review. Magnetic Resonance in Medicine, 69(3), 621–636. https://doi.org/10.1002/mrm.24314

Magland, J. F., & Childress, A. R. (2014). Task-Correlated Facial and Head Movements in Classifier-Based Real-Time fMRI. Journal of Neuroimaging, 24(4), 371–378. https://doi.org/10.1111/jon.12015

Menon, V., Lim, K. O., Anderson, J. H., Johnson, J., & Pfefferbaum, A. (1997). Design and efficacy of a head-coil bite bar for reducing movement-related artifacts during functional MRI scanning. Behavior Research Methods, Instruments, & Computers, 29(4), 589–594. https://doi.org/10.3758/BF03210613

Muraskin, J., Ooi, M. B., Goldman, R. I., Krueger, S., Thomas, W. J., Sajda, P., & Brown, T. R. (2013). Prospective active marker motion correction improves statistical power in BOLD fMRI. NeuroImage, 68, 154–161. https://doi.org/10.1016/j.neuroimage.2012.11.052

Murphy, W. J., Stephenson, M. R., Byrne, D. C., Witt, B., & Duran, J. (2011). Effects of training on hearing protector attenuation. Noise and Health, 13(51), 132. https://doi.org/10.4103/1463-1741.77215

OSHA. (2008). OSHA Regulations (Standards-29 CFR), Occupational noise exposure-1910.95. https://www.osha.gov/laws-regs/regulations/standardnumber/1910/1910.95

Pardoe, H. R., Kucharsky Hiess, R., & Kuzniecky, R. (2016). Motion and morphometry in clinical and nonclinical populations. NeuroImage, 135, 177–185. https://doi.org/10.1016/j.neuroimage.2016.05.005

Power, J. D., Barnes, K. a, Snyder, A. Z., Schlaggar, B. L., & Petersen, S. E. (2012). Spurious but systematic correlations in functional connectivity MRI networks arise from subject motion. NeuroImage, 59(3), 2142–2154. https://doi.org/10.1016/j.neuroimage.2011.10.018

Power, J. D., Mitra, A., Laumann, T. O., Snyder, A. Z., Schlaggar, B. L., & Petersen, S. E. (2014). Methods to detect, characterize, and remove motion artifact in resting state fMRI. NeuroImage, 84, 320–341. https://doi.org/10.1016/j.neuroimage.2013.08.048

Power, J. D., Silver, B. M., Silverman, M. R., Ajodan, E. L., Bos, D. J., & Jones, R. M. (2019). Customized head molds reduce motion during resting state fMRI scans. NeuroImage, 189(January), 141–149. https://doi.org/10.1016/j.neuroimage.2019.01.016

Pruim, R. H. R., Mennes, M., van Rooij, D., Llera, A., Buitelaar, J. K., & Beckmann, C. F. (2015). ICA-AROMA: A robust ICA-based strategy for removing motion artifacts from fMRI data. NeuroImage, 112, 267–277. https://doi.org/10.1016/j.neuroimage.2015.02.064

Pua, E. P. K., Barton, S., Williams, K., Craig, J. M., & Seal, M. L. (2020). Individualised MRI training for paediatric neuroimaging: A child-focused approach. Developmental Cognitive Neuroscience, 41, 100750. https://doi.org/10.1016/j.dcn.2019.100750

Ravicz, M. E., & Melcher, J. R. (2001). Isolating the auditory system from acoustic noise during functional magnetic resonance imaging: Examination of noise conduction through the ear canal, head, and body. The Journal of the Acoustical Society of America, 109(1), 216–231. https://doi.org/10.1121/1.1326083

Reuter, M., Tisdall, M. D., Qureshi, A., Buckner, R. L., van der Kouwe, A. J. W., & Fischl, B. (2015). Head motion during MRI acquisition reduces gray matter volume and thickness estimates. NeuroImage, 107, 107–115. https://doi.org/10.1016/j.neuroimage.2014.12.006

Rosen, A. F. G., Roalf, D. R., Ruparel, K., Blake, J., Seelaus, K., Villa, L. P., Ciric, R., Cook, P. A., Davatzikos, C., Elliott, M. A., Garcia de La Garza, A., Gennatas, E. D., Quarmley, M., Schmitt, J. E., Shinohara, R. T., Tisdall, M. D., Craddock, R. C., Gur, R. E., Gur, R. C., & Satterthwaite, T. D. (2018). Quantitative assessment of structural image quality. NeuroImage, 169, 407–418. https://doi.org/10.1016/j.neuroimage.2017.12.059

Satterthwaite, T. D., Wolf, D. H., Loughead, J., Ruparel, K., Elliott, M. a, Hakonarson, H., Gur, R. C., & Gur, R. E. (2012). Impact of in-scanner head motion on multiple measures of functional connectivity: Relevance for studies of neurodevelopment in youth. NeuroImage, 60(1), 623–632. https://doi.org/10.1016/j.neuroimage.2011.12.063

Savalia, N. K., Agres, P. F., Chan, M. Y., Feczko, E. J., Kennedy, K. M., & Wig, G. S. (2017). Motion-related artifacts in structural brain images revealed with independent estimates of in-scanner head motion. Human Brain Mapping, 38(1), 472–492. https://doi.org/10.1002/hbm.23397

Schellhammer, F., Ostermann, T., Krüger, G., Berger, B., & Heusser, P. (2013). Good scent in MRI: Can scent management optimize patient tolerance? Acta Radiologica, 54(7), 795–799. https://doi.org/10.1177/0284185113482606

Siegel, J. S., Mitra, A., Laumann, T. O., Seitzman, B. A., Raichle, M., Corbetta, M., & Snyder, A. Z. (2016). Data Quality Influences Observed Links Between Functional Connectivity and Behavior. Cerebral Cortex, 1–11. https://doi.org/10.1093/cercor/bhw253

Simhal, A. K., Filho, J. O. A., Segura, P., Cloud, J., Petkova, E., Gallagher, R., Castellanos, F. X., Colcombe, S., Milham, M. P., & Martino, A. D. (2021). Predicting multimodal MRI outcomes in children with neurodevelopmental conditions following MRI simulator training. BioRxiv, 2021.01.28.428697. https://doi.org/10.1101/2021.01.28.428697

Smith, S. M., Zhang, Y., Jenkinson, M., Chen, J., Matthews, P. M., Federico, A., & De Stefano, N. (2002). Accurate, Robust, and Automated Longitudinal and Cross-Sectional Brain Change Analysis. NeuroImage, 17(1), 479–489. https://doi.org/10.1006/nimg.2002.1040

Thesen, S., Heid, O., Mueller, E., & Schad, L. R. (2000). Prospective acquisition correction for head motion with image-based tracking for real-time fMRI. Magnetic Resonance in Medicine, 44(3), 457–465. https://doi.org/10.1002/1522-2594(200009)44:3<457::AID-MRM17>3.0.CO;2-R

Tisdall, M. D., Hess, A. T., Reuter, M., Meintjes, E. M., Fischl, B., & van der Kouwe, A. J. W. (2012). Volumetric navigators for prospective motion correction and selective reacquisition in neuroanatomical MRI. Magnetic Resonance in Medicine, 68(2), 389–399. https://doi.org/10.1002/mrm.23228

van der Zwaag, W., Francis, S., & Bowtell, R. (2006). Improved echo volumar imaging (EVI) for functional MRI. Magnetic Resonance in Medicine, 56(6), 1320–1327. https://doi.org/10.1002/mrm.21080

Van Dijk, K. R. a, Hedden, T., Venkataraman, A., Evans, K. C., Lazar, S. W., & Buckner, R. L. (2010). Intrinsic functional connectivity as a tool for human connectomics: Theory, properties, and optimization. Journal of Neurophysiology, 103(1), 297–321. https://doi.org/10.1152/jn.00783.2009

Van Dijk, K. R. A., Sabuncu, M. R., & Buckner, R. L. (2012). The influence of head motion on intrinsic functional connectivity MRI. NeuroImage, 59(1), 431–438. https://doi.org/10.1016/j.neuroimage.2011.07.044

Vanderwal, T., Kelly, C., Eilbott, J., Mayes, L. C., & Castellanos, F. X. (2015). Inscapes: A movie paradigm to improve compliance in functional magnetic resonance imaging. NeuroImage, 122, 222–232. https://doi.org/10.1016/j.neuroimage.2015.07.069

White, N., Roddey, C., Shankaranarayanan, A., Han, E., Rettmann, D., Santos, J., Kuperman, J., & Dale, A. (2010). PROMO: Real-time prospective motion correction in MRI using image-based tracking. Magnetic Resonance in Medicine, 63(1), 91–105. https://doi.org/10.1002/mrm.22176

Wong-Baker FACES Foundation. (2020). Wong-Baker FACES® Pain Rating Scale. Wong-Baker FACES Foundation. https://wongbakerfaces.org/

Wylie, G. R., Genova, H., DeLuca, J., Chiaravalloti, N., & Sumowski, J. F. (2014). Functional magnetic resonance imaging movers and shakers: Does subject-movement cause sampling bias? Human Brain Mapping, 35(1), 1–13. https://doi.org/10.1002/hbm.22150

Yan, C.-G., Cheung, B., Kelly, C., Colcombe, S., Craddock, R. C., Di Martino, A., Li, Q., Zuo, X.-N., Castellanos, F. X., & Milham, M. P. (2013). A comprehensive assessment of regional variation in the impact of head micromovements on functional connectomics. NeuroImage, 76, 183–201. https://doi.org/10.1016/j.neuroimage.2013.03.004

Yang, S., Ross, T. J., Zhang, Y., Stein, E. A., & Yang, Y. (2005). Head motion suppression using real-time feedback of motion information and its effects on task performance in fMRI. NeuroImage, 27(1), 153–162. https://doi.org/10.1016/j.neuroimage.2005.02.050

Yendiki, A., Koldewyn, K., Kakunoori, S., Kanwisher, N., & Fischl, B. (2014). Spurious group differences due to head motion in a diffusion MRI study. NeuroImage, 88, 79–90. https://doi.org/10.1016/j.neuroimage.2013.11.027

Zaitsev, M., Dold, C., Sakas, G., Hennig, J., & Speck, O. (2006). Magnetic resonance imaging of freely moving objects: Prospective real-time motion correction using an external optical motion tracking system. NeuroImage, 31(3), 1038–1050. https://doi.org/10.1016/j.neuroimage.2006.01.039

Zaitsev, Maxim, Maclaren, J., & Herbst, M. (2015). Motion artifacts in MRI: A complex problem with many partial solutions. Journal of Magnetic Resonance Imaging, 42(4), 887–901. https://doi.org/10.1002/jmri.24850

